# Reconciling heterogeneous dengue virus infection risk estimates from different study designs

**DOI:** 10.1101/2024.09.09.24313375

**Authors:** Angkana T. Huang, Darunee Buddhari, Surachai Kaewhiran, Sopon Iamsirithaworn, Direk Khampaen, Aaron Farmer, Stefan Fernandez, Stephen J. Thomas, Isabel Rodriguez Barraquer, Taweewun Hunsawong, Anon Srikiatkhachorn, Gabriel Ribeiro dos Santos, Megan O’Driscoll, Marco Hamins-Puertolas, Timothy Endy, Alan L. Rothman, Derek A. T. Cummings, Kathryn Anderson, Henrik Salje

## Abstract

Uncovering rates at which susceptible individuals become infected with a pathogen, i.e. the force of infection (FOI), is essential for assessing transmission risk and reconstructing distribution of immunity in a population. For dengue, reconstructing exposure and susceptibility statuses from the measured FOI is of particular significance as prior exposure is a strong risk factor for severe disease. FOI can be measured via many study designs. Longitudinal serology are considered gold standard measurements, as they directly track the transition of seronegative individuals to seropositive due to incident infections (seroincidence). Cross-sectional serology can provide estimates of FOI by contrasting seroprevalence across ages. Age of reported cases can also be used to infer FOI. Agreement of these measurements, however, have not been assessed. Using 26 years of data from cohort studies and hospital-attended cases from Kamphaeng Phet province, Thailand, we found FOI estimates from the three sources to be highly inconsistent. Annual FOI estimates from seroincidence was 2.46 to 4.33-times higher than case-derived FOI. Correlation between seroprevalence-derived and case-derived FOI was moderate (correlation coefficient=0.46) and no systematic bias. Through extensive simulations and theoretical analysis, we show that incongruences between methods can result from failing to account for dengue antibody kinetics, assay noise, and heterogeneity in FOI across ages. Extending standard inference models to include these processes reconciled the FOI and susceptibility estimates. Our results highlight the importance of comparing inferences across multiple data types to uncover additional insights not attainable through a single data type/analysis.

**Significance statement:** Dengue virus infections are surging globally. Knowing who, where, and how many people are at risk of infection is crucial in determining means to protect them. Here, we compare three current approaches in measuring risk (two involving blood samples and one involving case counts) to estimate the risk of infection. Estimates derived from each method differed greatly. By accounting for rise and falls of antibodies following infections, noise in the antibody titer measurements, and heterogeneity in infection risk across ages, we reconciled the measurements. As measurements from blood samples and case counts are pillars in uncovering risk of most infectious diseases, our results signifies integrating these processes into risk measurements of pathogens beyond dengue virus.

## Introduction

Quantifying historical infection intensity of pathogens is essential to assess infection burden and susceptibility of populations through time, insights that are pivotal in predicting future transmission potentials and shaping effective intervention strategies (1–3). Infection intensity is often quantified as the rate at which susceptible individuals become infected, a concept known as the Force of Infection (FOI). For dengue virus (DENV) infections, quantifying infection risk through FOI is of particular significance, as infection burden is non-linearly linked to the observable disease burden First infection by one of the four DENV serotypes is primarily subclinical but the generated immune response is the most widely recognized risk factor for severe disease following a second infection by a different serotype (4). The FOI can also be used to estimate how immunity is distributed in the population (by age, for example) to identify who is at risk of infections having already acquired immunity (5–7). Information on infection risk in populations and the distribution of immunity are integral to optimizing the impact of the two currently licensed vaccines and avoiding deleterious outcomes (8–14).

Typically, two main sources of data are employed to estimate historical infection intensity, or FOI, in populations: serological data and case count data. In parallel, two different study designs have been used to estimate forces of infection: longitudinal and cross-sectional. Longitudinal serological studies are often considered the gold standard, as they directly track the transition of seronegative individuals to seropositive (seroincidence) (7, 15). Cross-sectional serological data, which includes individuals of different ages, can provide estimates of FOI by drawing upon differences in exposure histories across birth cohorts (16–20). Similarly, age-stratified case count data can extract information from age distribution of cases over time which reflects the variation in exposure histories among different age groups (21–24). To infer FOI from age-stratified case counts, models are employed to link the infection process with the generation of reported cases. The model typically accounts for reporting rates, but can also include processes that influence illness manifestations (23).

These approaches rely on different assumptions about antibody responses following infection, age-specific differences in infection risk, the role of cross-reactivity from infection or vaccination from related viruses, accuracy of the serological assay, and how immunity preceding infections affects the risk of symptoms. However, the importance of these different assumptions on the resulting FOI estimates is largely unknown. Further, little is known about the consistency in estimates derived from the different approaches. In this study, we leverage 26 years of data from a single location to compare FOI estimates obtained from various data types. In this single location both serological and clinical case data is available from longitudinal cohorts and from a passive surveillance system. We compare estimates derived from different subsets of the available data, identify the sources of discrepancies and develop methods to improve estimates through joint inference when multiple data types are available.

## Results

### Dengue data in Kamphaeng Phet

Kamphaeng Phet province, Thailand (KPP) represents a dengue hyper-endemic region with four consecutive longitudinal cohort studies conducted from 1998 to the present: Kamphaeng 42 Phet Prospective Study 1 (KPS1, 1998-2002), KPS2 (2004-2007), KPS3 (2010), and Kamphaeng Phet Family Cohort Study (KFCS, 2015-ongoing) (25–28), **Figure 1** and **Table S1**. KPS1 and KPS2 were school children cohorts while KPS3 was a one-year cohort of children in the community. KFCS is a community cohort focused on multi-generational households (28). Individuals were bled every 3, 6, 6, and 12 months in these cohorts, respectively, and tested for anti-DENV antibodies via hemagglutination inhibition assay (HAI). Percentages of seropositive samples (geometric mean titer (GMT) >=10) increased with age except for samples obtained at very young ages, attributable to the presence of maternally-derived antibodies and cross-reactive antibodies from Japanese Encephalitis vaccination (29, 30), **Figure 1b**. Among participants aged nine, 75%, 57%, 53%, and 49% have GMT>=10, respectively. All individuals in KFCS have GMT>=10 after age 30 (97% with GMT>=20).

**Figure 1.**
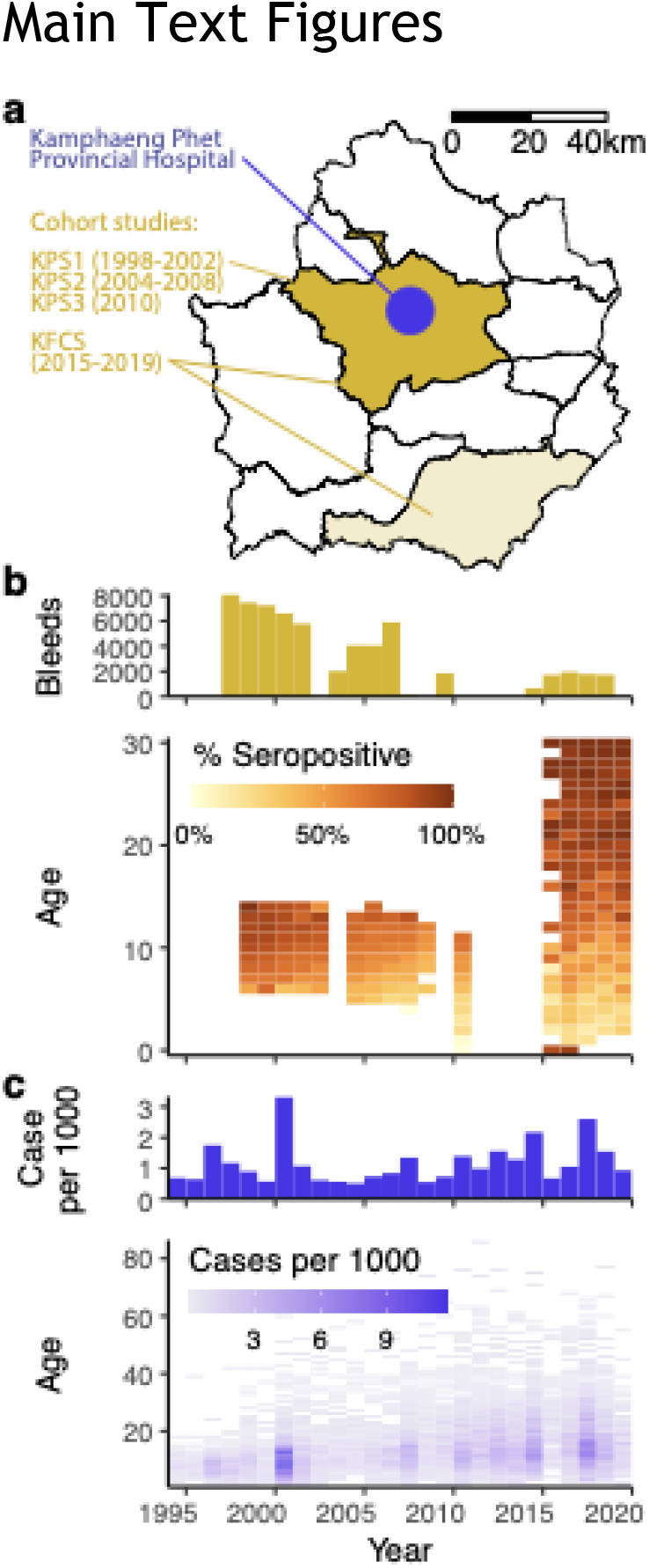
Study data. **a)** Map of Kamphaeng Phet province showing spatial coverage of cohort studies (colored) and location of Kamphaeng Phet Provincial Hospital (KPPH, blue point). **b)** Number of bleeds by year (top) and percentages of with GMT>=10 by age and year of collection (bottom). **c)** Number of dengue cases reported at KPPH per thousand population by year (top), and by year and age (bottom).

Within Mueng, the capital district, the Kamphaeng Phet Provincial Hospital (KPPH) serves as the sole tertiary care facility in the province. Between 1994 and 2020, KPPH reported a total of 17,773 cases suspected of dengue among KPP residents (of which 12,819 were lab confirmed), representing an annual incidence of 0.5 to 3.3 cases per thousand population (**Figure 1c**). Mueng residents accounted for 55% of these cases.

### Inferred force of infection (FOI) differs across data sources

Considering the cohorts as both longitudinal measures (multiple samples per individual) and cross-sectional data (single sample per individual), we estimated the annual per-serotype FOI between 1998 and 2019 using standard models for each data type (16–20), **Figure S1**. Bleeds taken before age three were excluded to avoid interference from maternally-derived antibodies and/or cross-reactive antibodies from Japanese Encephalitis vaccination. We derived case-based FOI by fitting a model which takes into account differences in symptomatic rates across the four possible infections of individuals (one by each serotype) and variations in time and age for DENV-infected individuals to seek care at KPPH (23). We excluded cases under age one as their symptomatic rate upon first DENV infection differs from the others due to maternally-derived immune-enhancement (31). All models assumed that infection risks in the excluded ages remained similar to the rest of the population despite differences in test positive tendencies or clinical presentations.

Applying a standard geometric mean titer (GMT) threshold of 10 to define seropositivity of serological samples, we found that the seroincidence-derived annual FOIs were consistently higher than the case-derived annual FOIs (3.40-fold on average, 95%CI: 2.46 to 4.33, **Figure 2a**) and seroprevalence-derived annual FOIs (95%CI: 1.70, 3.96-folds). Ratios between cross-sectional seroprevalence-derived annual FOIs and case-derived annual FOIs did not appear to vary systematically (95%CI: 0.72, 1.23-folds), with moderate correlation between the two (correlation coefficient=0.46). The estimates derived from both serological sources exhibited wide uncertainty. Raising the GMT threshold to 20 to mitigate false positives from e.g., individuals seropositive from JEV vaccination, did not lower FOI estimates from seroincidence (95%CI: 1.95, 3.55-folds). However, it led to systematically lower seroprevalence-derived FOI relative to case-derived FOI (0.43 to 0.81-fold, **Figure S2a**). The discrepancy patterns remained similar when compared to case-derived FOIs inferred from lab-confirmed cases (**Figure S3**).

**Figure 2.**
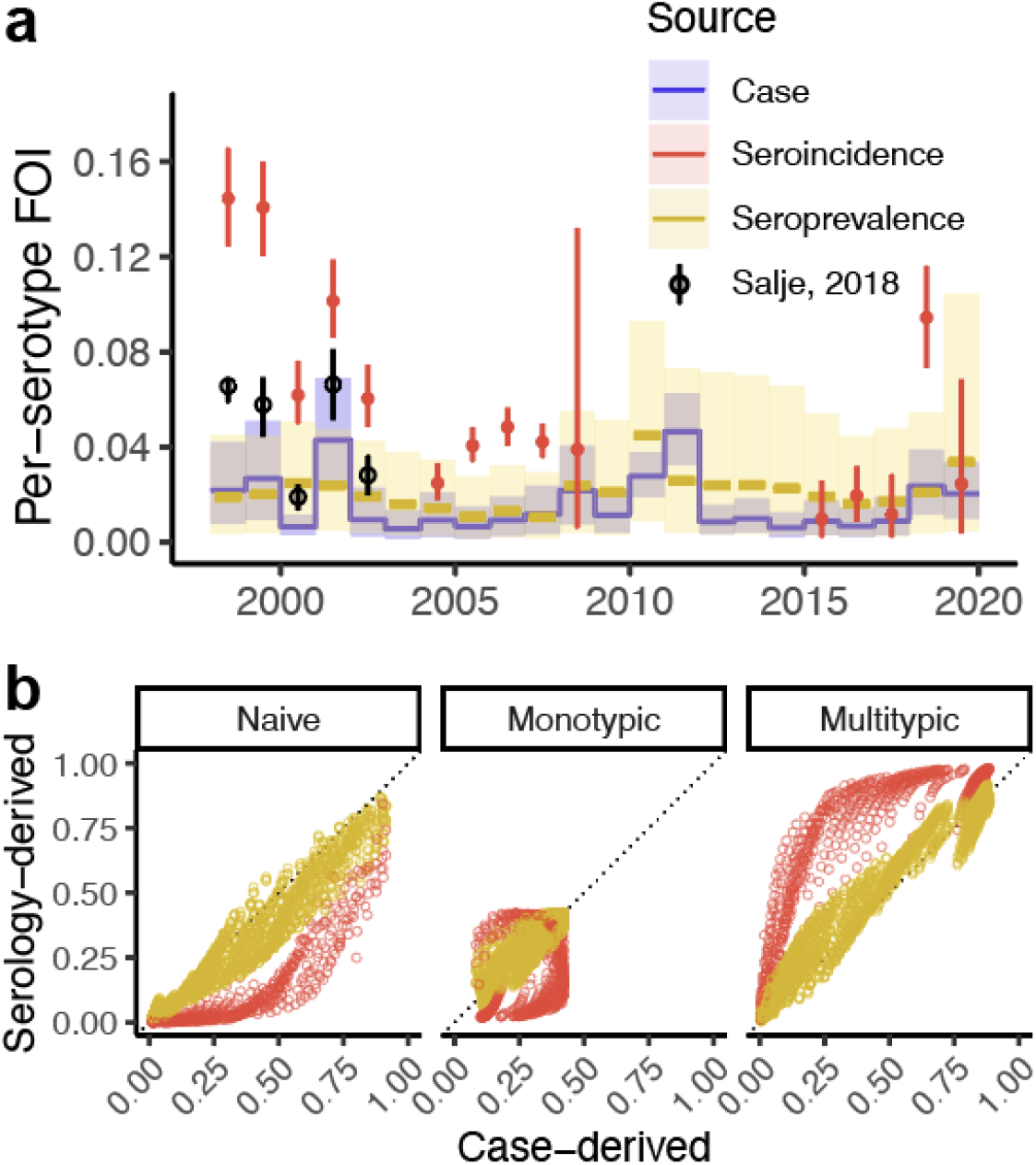
Estimates from standard force of infection (FOI) inference models. **a)** Annual FOI estimated from each of the data sources: seroincidence data (red) and seroprevalence data (yellow) using seropositivity threshold of GMT>=10, and case data (blue). Annual FOI estimated from longitudinal samples of KPS1 (black, (7)) are included for comparison. **b)** Relationship between serology-derived (y-axis, respective colors) and case-derived susceptibility reconstructions (x-axis). Each point in the reconstruction represents the proportion in each age-year that has not been infected with DENV (naive), has been infected by one serotype (monotypic) or more than one serotype (multitypic).

These discrepancies and uncertainties in FOI values resulted in notable differences in the reconstructed susceptibility fractions across the different approaches (see **Figure 2b**). For instance, in the most recent year of the study (2019), case-derived reconstructions suggested 56% of 9yrs old remained DENV-naive (95%CI: 49%, 64%) while seroprevalence-derived and seroincidence-derived reconstructions suggested 40% (95%CI: 30%, 51%) and 24% (95%CI: 12%, 37%) of 9yrs old remained DENV-naive, respectively.

### Simulations to study effects of violated model assumptions on inferred FOI

To identify sources of the FOI discordance, we performed an extensive suite of simulations in which data generation and true infection rates were known. We analyzed simulated data using our different approaches described above. Our simulations incorporated varying assumptions of the effects of waning antibody titers, measurement error in assay readouts, and titers against cross-reactive pathogens. Prior research has demonstrated that following primary DENV infections, antibody titers rise rapidly but then wane exponentially to a steady titer approximately 5-times lower within a year (7). After a subsequent infection by a different DENV serotype, titers increase to levels that are robust to detection. Measurement error in assay readouts can lead to titers falling below seropositivity thresholds, while individuals without prior exposure to DENV may exhibit seropositivity due to titers against other flaviviruses (29, 32), **Figure 3a**. Additionally, variations in infection risk across different age groups are possible (23). We simulated infection timings of 500,000 individuals with defined FOI by year to eliminate imprecisions in estimates resulting from insufficient statistical power to study effects of these processes on the inferred FOI.

**Figure 3.**
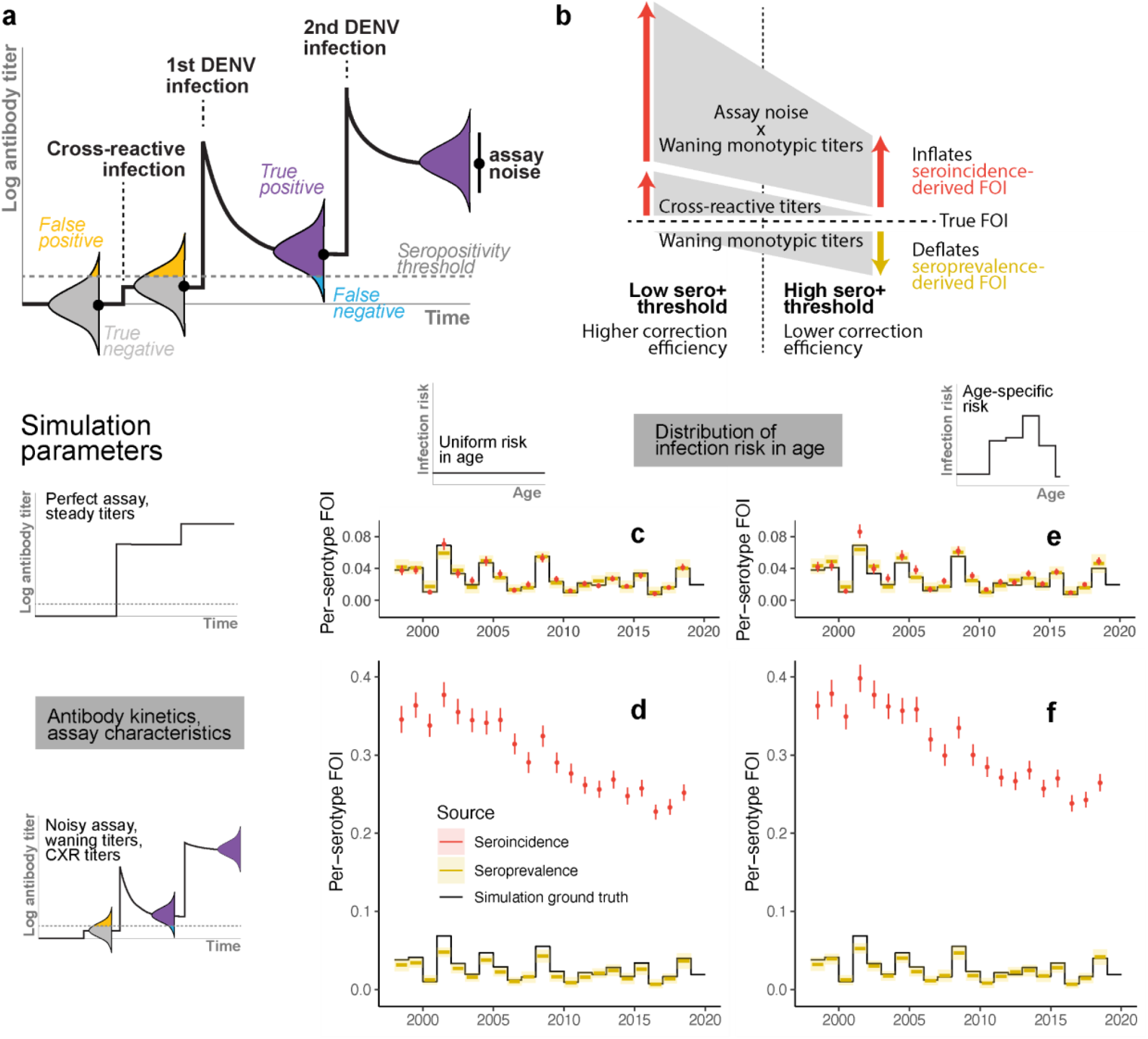
Biases in serology-derived force of infection (FOI) using simulated data with known true parameters. **a)** Illustration of anti-DENV antibody kinetics as an individual acquires a cross-reactive (CXR) virus infection or vaccination (i.e., not DENV), one DENV infection, and >1 DENV infections. Measured titers distribute around the true underlying titers with variability depending on the assay characteristics. **b)** Schematic of biases in serology-derived FOI and their correction efficiencies at low and high seropositivity thresholds. **c-f)** Antibody kinetics, assay characteristics (rows), and distribution of infection risk in age among susceptible individuals (columns) used to generate observed titer measurements and FOI inferred from those respective simulations using standard models for seroincidence (red) and seroprevalence (yellow).

We found that in the absence of random measurement error and when the seropositivity threshold is low enough to correctly discriminate DENV-exposed individuals from naives, waning monotypic titers do not lead to biased FOI estimates from either serological data types (**Figure S4a-b** and **Figure S5a**). However, when the simulation included random measurement error, using a low threshold led to false positives which inflated seroincidence-derived FOI (**Figure S4c**). The inflation was exacerbated by the presence of cross-reactive titers (**Figure 3d, Figure S4d**). While raising the threshold to define seropositivity helped mitigate inflation if titers of exposed individuals remained high (**Figure S5d**), the trade-off for false negatives when monotypic titers did wane led to even more pronounced over-estimates of FOIs (**Figure S5e-f**). The over-estimation arose from the greater chance of testing positive in the follow-up bleed in DENV-exposed individuals that tested falsely negative at pre-interval compared to truly DENV-naive individuals. In fact, the over-estimation can be severe even at lower thresholds where fewer false negative individuals were expected (**Figure S4e-f**, see also **Supplementary Mathematical Analysis)**.

In the absence of waning titers, seroprevalence-derived FOI appeared robust to assay noise and cross-reactive titers at both seropositivity thresholds (**Figure S4c-d, Figure S5c-d**). Expectedly, false negativity due to waning monotypic titers led to underestimation of FOI, which became more extreme with higher seropositivity thresholds (**Figure S4e-f, Figure S5e-f**).

We found that age-specific differences in risk of infection could also cause discrepancies in FOI estimates. Even when the susceptibility status of individuals could be perfectly ascertained, seroincidence-derived FOI estimates were systematically different from seroprevalence-derived FOI. (**Figure 4e, Figure S4g, Figure S5g**). When age-specific risk was present in conjunction with waning titers and assay noise, the difference was exacerbated (**Figure 3f**).

**Figure 4.**
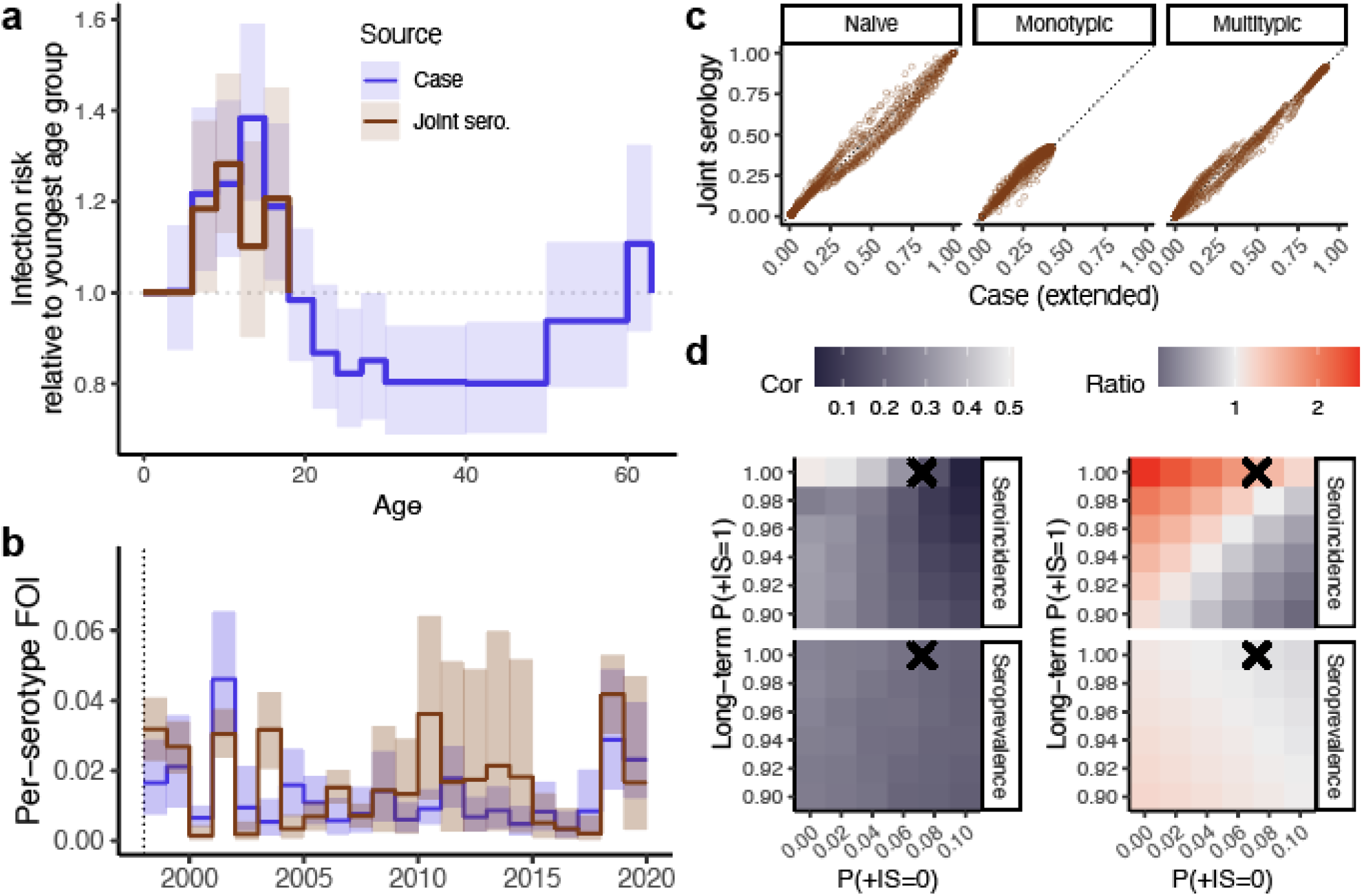
Infection risks in Kamphaeng Phet. **a)** Distribution of infection risk by age, **b)** annual per-serotype FOI inferred from the joint serology model (brown) and the extended case-based model (blue), and **c)** relationships between the susceptibility reconstructions. Each point in the reconstruction represents the proportion in each age-year that has not been infected with DENV (naive), has been infected by one serotype (monotypic) or more than one serotype (multitypic). **d)** Effects of presumed test positive probabilities in DENV-naives (x-axis) and long-term test positive probabilities in monotypically-infected individuals (y-axis) on the correlation and ratio between temporal FOIs inferred from the extended case-based model and temporal FOIs inferred from a single data source (either seroincidence or seroprevalence at seropositivity threshold of 10) imposed with age-specific risk inferred from the extended case-based model. Test positive probabilities estimated from the joint serology model are annotated as crosses for comparison.

### Correcting for violated assumptions recovers temporal FOI of simulation ground truth at varying efficiencies

In highly powered datasets, when only assay noise was present, we found that inflation in estimated seroincidence could be perfectly mitigated at both seropositivity thresholds (10 and 20, **Figure S6a, Figure S7a**) by correcting for (presumed known) test positive probabilities given the susceptibility status of individuals and the distribution in infection risk by age. In the presence of both assay noise and waning monotypic titers, this simple correction, which did not take into account exact infection timings of individuals and the variable amounts of titers waned across individuals, reduced but did not eliminate the inflation in estimated seroincidence-derived FOI (**Figure S6b, Figure S7b**). Importantly, the new estimates at a threshold of 20 showed greater discrepancies from the ground truth than at a threshold of 10 (1.35 to 1.63-fold difference as compared to the ground truth vs 0.81 to 0.98-fold difference). The correction efficiencies remained similar when additional corrections for cross-reactive titers in DENV-naives and non-uniform risk in age were needed (**Figure S6c-d, Figure S7c-d**). In contrast, the simple correction was able to efficiently correct for biases in seroprevalence-derived FOI in all cases (**Figure S6a-e, Figure S7a-e**).

When we reduced the size of the simulated datasets to match the number and time points of samples available in our cohort studies, we found that correlation with the ground truth for both serological data types was lower despite the same adjustments to correct for assay noise, waning monotypic titers and age-specific differences in risk (**Figure S6g** vs **Figure S6d**). Uncertainties in the estimates increased substantially (**Figure S6f-g**).

### Reconciled infection risk in KPP is non-uniform across ages

Taking into account sources of discrepancies in estimation of FOI learnt from the simulation studies, namely, age-specific infection risk, antibody kinetics, and assay variability, we developed a model that is jointly informed by both serological data types to estimate the shared underlying infection risk in KPP and a separate case-based model with age-specific risk extension.

Infection risk estimates from the ‘joint serology model’ and the extended case-based model showed good agreement, **Figure 4a-b**. Both models suggested elevated infection risk in KPP between ages 6-17yrs compared to the reference class (age 0-5yrs). Correlation between the temporal FOIs was moderate (cor. coef.=0.48) but without systematic differences in magnitudes (ratio between serology-based to cased-based FOIs of 0.72 to 1.34). Reconstructions of susceptibility by age and year from the two models were highly congruent (cor. coef>=0.97 without signs of systematic differences, **Figure 4c**).

### Test positive probabilities are key to the reconciliation

Using multiple serological data sources with shared underlying processes, we were able to characterize factors governing probabilities of falsely testing positive at various thresholds when DENV-naive, and testing positive when DENV-exposed, in tandem with the infection risks. The factors are namely the post-infection antibody titers captured in the serological samples and variability in the assay measurements, **Figure S10**. We estimated that the captured titer rises between bleeding intervals of the cohort study participants in response to primary DENV infections that occurred during the intervals were comparable across studies: an average rise of 7.89 log2 (95%CI: 4.83, 13.10). The titers then declined to a steady level of 2.76 log2 (95%CI: 2.53, 2.96). Standard deviation of assay readouts was estimated to be 0.51 (95%CI: 0.36, 0.64) which corresponds to 99.9% to 100% of monotypic titers above a threshold of 10 over the long-term (i.e., cross-sectional seroprevalence studies) or 92.7% to 93.7% at a threshold of 20. Averaged across the studies, DENV-naive individuals have a 6.2% to 7.3% chance of testing positive for DENV at threshold of 10 and <0.2% at threshold of 20.

We re-inferred temporal FOIs from each of the serological data sources presuming various other sets of test positive probabilities and found that congruence with case-derived FOIs were reduced (**Figure 4d, Figure S11**). Importantly, FOIs from seroincidence varied greatly across test positive probabilities leading to pronounced changes in congruence compared to seroprevalence-derived FOIs.

## Discussion

Leveraging a unique opportunity where over two decades of longitudinal serological data and hospital case count data are available from the same community, we assessed the congruence in FOI estimated from different data types. We found large discrepancies between the FOI estimates. Consequently, susceptibility in the population inferred from the estimates were drastically different. Our investigations revealed causes of these discrepancies as a lack of accounting for antibody kinetics and assay noise (affecting serological data types), and age-specific infection risk (affecting all data types).

Longitudinal serology is crucial to track infections of individuals and ascertain their evolving exposure statuses (5, 7). However, we found identifying DENV infections based on seroconversion from negative to positive at a specified threshold was highly sensitive to the interplay between antibody kinetics and assay noise. Importantly, the heterogeneous seroconversion tendencies across sera pairs could not be efficiently corrected for by applying average test positive probabilities across sera pairs. Our findings support the use of individual-based titer reconstructions, mechanistically taking into account sources of bias, to detect infections (7, 33, 34).

Cross-sectional serology is the most common data source used to infer dengue burden (17, 19, 35–38). While this approach bypasses biases in case reporting, our findings highlight that the processes that affect a test coming back positive or not should be considered (39). Applying a single seroreversion rate to all exposed individuals may be sufficient to account for waning antibodies in non-endemic settings. However, given the increased durability of antibodies in multitypically exposed individuals (40), settings with multiple serotypes co-circulating will need to account for heterogeneity in antibody kinetics across individuals with different infection histories. The likely exposure to cross-reacting viruses (e.g., JEV or ZIKV) due to shared vector ecology or vaccination will also require appropriate corrections. Our analyses assumed that cross-reactive titers were evenly present in DENV-naives aged >=3yrs. This may be true for Japanese Encephalitis vaccine-induced titers as children were vaccinated widely at young ages (<=2yr (41)) but would only be true for co-circulating viruses such as ZIKV (29, 42) if FOI of these viruses were extremely high such that all children were exposed before age 3. Further explorations are needed to assess how heterogeneous presence of cross-reactive titers would impact estimation of FOI of DENV, especially if these exposures alter antibody kinetics to subsequent DENV infections (43).

Age-stratified case data provides an alternative means to estimate past dengue burdens (21– 23). As the method involves adjusting for multiple processes to uncover the true age distribution of infections (which is informative of the infection burden) from the observed age distribution, long periods of surveillance are necessary to reliably estimate FOI. With 26 years of data fine-scale age strata, our FOI estimates from case data tracked closely with estimates from our joint model (our best approximation of the underlying dengue burden). In settings with less data, the ability to disentangle observation processes from infection risk would be more limited making simplifying assumptions necessary. Analytical studies and simulations are needed to assess the impact of those assumptions on the inferred FOI.

Our inferences revealed evidence of age-specific infection risk, a characteristic often neglected in dengue epidemiology. Whether the variation reflects behavioral, immunological, and/or physiological differences (44–46), the heterogeneity challenges generalizations of risk measured in a sample to the general population. Uncovering processes leading to these differences is key to overcoming this challenge and will facilitate the development and management of interventions.

The importance of characterizing antibody kinetics, assay variability, and distribution of risk in the population demonstrated in our study applies broadly beyond dengue as serological and case data are pillars in quantifying infection burdens in most diseases. Our results highlight the need to compare inferences across multiple data types and analysis methods both to flag blindspots in each of the inferences to mitigate misconclusions and to uncover additional insights not attainable through a single data type/analysis.

## Material and methods

### Ethics statement

Use of data from the Kamphaeng Phet Hospital were reviewed and approved by Walter Reed Army Institute of Research Institutional Review Board (protocol number 1313 and 1957). The study protocol for KPS1 was approved by the Office of the Army Surgeon General, University of the Massachusetts Medical School, and the Ministry of Public Health, Thailand. The protocol for KPS2 was additionally approved by the University of California–Davis and San Diego State University (protocol number 654 and 1042). The KPS 3 and KFCS cohort study was approved by the Thailand Ministry of Public Health Ethical Research Committee; Siriraj Ethics Committee on Research Involving Human Subjects; Institutional Review Board for the Protection of Human Subjects, State University of New York Upstate Medical University; and Walter Reed Army Institute of Research Institutional Review Board (protocol number 1552 and 2119).

### Empirical data for serological models

Longitudinal samples from four longitudinal cohort studies in Kamphaeng Phet province were included in this study: KPS1 (1998-2002), KPS2 (2004-2008), KPS3 (2010), and KFCS (2016-2019) (27). KPS1, 2 and 3 were cohorts of school children while KFCS focused on multi-generational households. To generate cross-sectional seroprevalence data from the longitudinal samples, one sample was randomly selected per individual. In the family cohort study, KFCS, only one randomly selected individual was selected per family to avoid reported interdependence between family members (27). Inferences involving seroprevalence data were repeated using three independent random samples.

Antibody titers were measured using hemagglutination inhibition assay (HAI) against DENV1 (Hawaii strain), DENV2 (New Guinea C strain), DENV3 (H87 strain), and DENV4 (814669 strain in KPS1-3 and H241 strain in KFCS) as described elsewhere (46, 47). Measurements were done in 2-fold serial dilutions between 1:10 and 1:10240. Titers <10 and >10240 were imputed as 5 and 20480, respectively. For each sample, geometric mean titers (GMT) were computed from the four serotype-specific titers. The linear scale GMT, A_linear_, were log transformed via equation log_2_(A_linear_ ⁄ 10) + 1 so that samples with linear scale titers of <10 to all four serotypes were zero.

### Empirical data for case-based models

Age-annotated cases suspected for dengue that sought care at the Kamphaeng Phet Provincial Hospital (KPPH) between 1994 and 2020 were included in this study. Cases were considered lab-confirmed when acute samples from the patients were tested positive for DENV via polymerase chain reaction (PCR) or virus isolation, or enzyme-linked immunosorbent assays (ELISAs) as per criteria described elsewhere (48–50). Inferences were restricted to Mueng residents to match the spatial coverage of the cohort studies. Population age censuses for 1994-2020 were acquired from the Department of Provincial Administration, Ministry of the Interior through the Official Statistics Registration Systems (51).

### Simulated data

We simulated one million observations (two bleeds of three months apart per individual for 500,000 individuals) to eliminate imprecisions of estimates from insufficient power. Observations were made between ages 5-15 years where the occurrence of first infections was expected to be concentrated. First, we simulated ages at which the individuals acquired their first, second, third, and fourth DENV infections under defined annual (and age-specific) force of infections. We then used anti-DENV antibody kinetics reported in Salje, 2018 (6) to generate true underlying titers of individuals at the observation time points. For scenarios where assays were imperfect, the observed titers were drawn from normal distributions with means equal to the true underlying titers and standard deviation σ = 0.49. In scenarios where non-zero titers in DENV-naives due to the presence of cross-reactive titers were considered, titers in DENV-naive individuals were drawn from a normal distribution with mean 0.266 (20% of the long-term titer in monotypic sera) and standard deviation σ_0_ = σ = 0.49.

To simulate data with power equivalent to empirical serological data, we performed the same procedures but with the number of individuals, observation time points and ages matching those in the cohort studies.

### Standard models in FOI inferences

Inferring force of infection (FOI, λ) of dengue from non-serotype-specific datasets typically assumes equivalent FOI across all serotypes in circulation, long-lived protection against the infecting serotype, and no cross protection against other serotypes, **Figure S1**. Hence, probability that an individual birth cohort *h* has escaped a particular serotype up to age *a* is 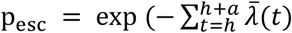 where 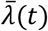 is the average per-serotype force of infection at time *t*. It follows that the probability that the individual has acquired *i* infections is

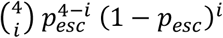

Assuming that antibodies in infected individuals are robustly above a chosen positivity threshold, FOI can be linked to **cross-sectional serology** via probability of testing positive written as 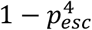 In **longitudinal serological studies**, the probability that a seronaive individual at time *t* tests positive at *t* + Δ*t* is similarly 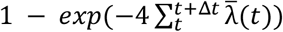

Increase in the proportion of individuals in birth cohort *h* that have acquired at least *i* infections between time *t* and *t* + Δ*t* indicates occurrence of the i-th infection during the time interval, *I*_Δ*t*_(*i, h, t*). Let proportion *Q*(*i*) of the i-th infections of individuals result in clinical presentations that are severe enough to trigger care seeking and proportion ϕ(*a, t*) of those severe infections go on to report to KPPH, *a* being the age of cohort *h* at time *t*. We express ϕ(*a, t*) as ϕ(*a*) · ϕ(*t*) where ϕ(*a*) is the piecewise constant age-specific reporting rate and ϕ(*t*) is the piecewise constant time-specific reporting rate. Considering *Pop* (*h, t*) the population size of birth cohort *h* at time *t*, we would expect the number of dengue cases from this birth cohort in this time interval who reported to KPPH to be

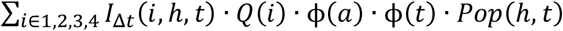

For **case counts aggregated by age**, the expected counts is the sum of expected cases of those birth cohorts contributing to the respective age bins.

### Extended models for FOI inferences

Standard models for all data types were extended to estimate age-specific infection risks relative to the youngest age group. Age groups were defined with consideration of data points available to inform estimates for the groups and consistency between data types to ease comparison, see **Table S5**.

The **joint serology model** estimates parameters characterizing antibody kinetics and assay noise in tandem with the infection risks. Probabilities of testing positive given susceptibility states of individuals were then derived from those parameters. Due to known significant waning in monotypic titers, we allowed for differing probabilities of testing positive in pre-vs post-interval bleeds for individuals who have been infected once to reflect their difference in expected time since infection. To derive these probabilities, we estimate the level of long-lasting titers Ω_*long*_ present in both bleeds and the additional short-lived titers Ω_*short*_ present only in post-interval bleeds (see **Figure S10**). Cross-reactive titers in DENV-naives, shared across studies, were estimated relative to Ω_*long*_. Following Salje et al (6), we formulated the relationship between true underlying titers of individuals *A*, standard deviation of assay measurement around the true titer σ, and probability of testing positive *P*(⊕ |*A*, σ) as 1 ‐ Φ((*A* ‐ *v*)⁄σ) where Ф is the cumulative density function of a standard normal distribution. Because bleeding intervals differed across cohort studies, we allowed Ω_*short*_ to differ across studies. The test positive probability for seroprevalence data was assumed to be the same as pre-interval bleeds. Likelihood was evaluated against seroprevalence and seroincidence data at two seropositivity thresholds (10 and 20) to better inform this relationship.

### Model fitting

In all models, we estimate annual FOI from ten years prior to the first observation to the year of the last data point. FOI prior to the ten years were assumed to be constant. We used Bernoulli likelihood to fit to serological data and negative binomial likelihood to fit to case data with priors as defined in **Table S2**.

Parameters of all models were estimated from the data using Rstan v2.21.2 (52) with five independent chains, each of length 2,000 (200 discarded as warm-up). Posteriors of all chains combined were considered converged when R-hat < 1.1 and effective sample size > 300 for all parameters. Where inferences were done for three repeated random samples (i.e. models involving seroprevalence data), reported parameter estimates were from posterior draws pooled across the repeats. Convergence was assessed prior to the pooling.

### Quantifying congruence

We quantify congruence between any two sets of estimates via Pearson’s correlation and an average ratio between their posterior medians. The average ratio was obtained by fitting a linear regression with zero intercept between the posterior medians and 95% confidence interval of the ratio was calculated as the point estimate +/-1.96 * standard error.

## Supporting information

Supplementary Mathematical Analysis

## Data Availability

Data and code used to generate all results are available at https://zenodo.org/doi/10.5281/zenodo.11635046.

## Data and code availability

Data and code used to generate all results are available at 350 https://zenodo.org/doi/10.5281/zenodo.11635046.

## Acknowledgement

This study was funded by the Military Infectious Disease Research Program and the US National Institutes of Health (program project award P01 AI034533 and 1R01AI175941-01). ATH was supported by the Herchel Smith Fellowship.

## Disclaimer

Material has been reviewed by the Walter Reed Army Institute of Research. There is no objection to its presentation and/or publication. The opinions or assertions contained herein are the private views of the author, and are not to be construed as official, or as reflecting true views of the Department of the Army or the Department of Defense. The investigators have adhered to the policies for protection of human subjects as prescribed in AR 70–25.

