## Supplementary Figures and Tables for "Reconciling heterogeneous dengue virus infection risk estimates from different study designs"

#### Data descriptions

**Table S1. Descriptions of serological data included in this study.**

| **Cohort study**<br>(study population) | **Years** | **Ages** | **N** |
|---|---|---|---|
| **KPS1**<br>(Primary school children) | 1998 to 2002 | 4 to 16 | 35,213 blood samples<br>3,436 individuals |
| **KPS2**<br>(Primary school children) | 2004 to 2008 | 3 to 15 | 3,180 blood samples<br>16,201 individuals |
| **KPS3**<br>(Community children) | 2010 | 3 to 11 | 1,659 blood samples<br>16,201 individuals |
| **KFCS**<br>(Mother-infant and their multigenerational family members) | 2016 to 2019 | 3 to 30 | 3,491 blood samples<br>1,116 individuals<br>318 family clusters |

#### Standard models in FOI inferences

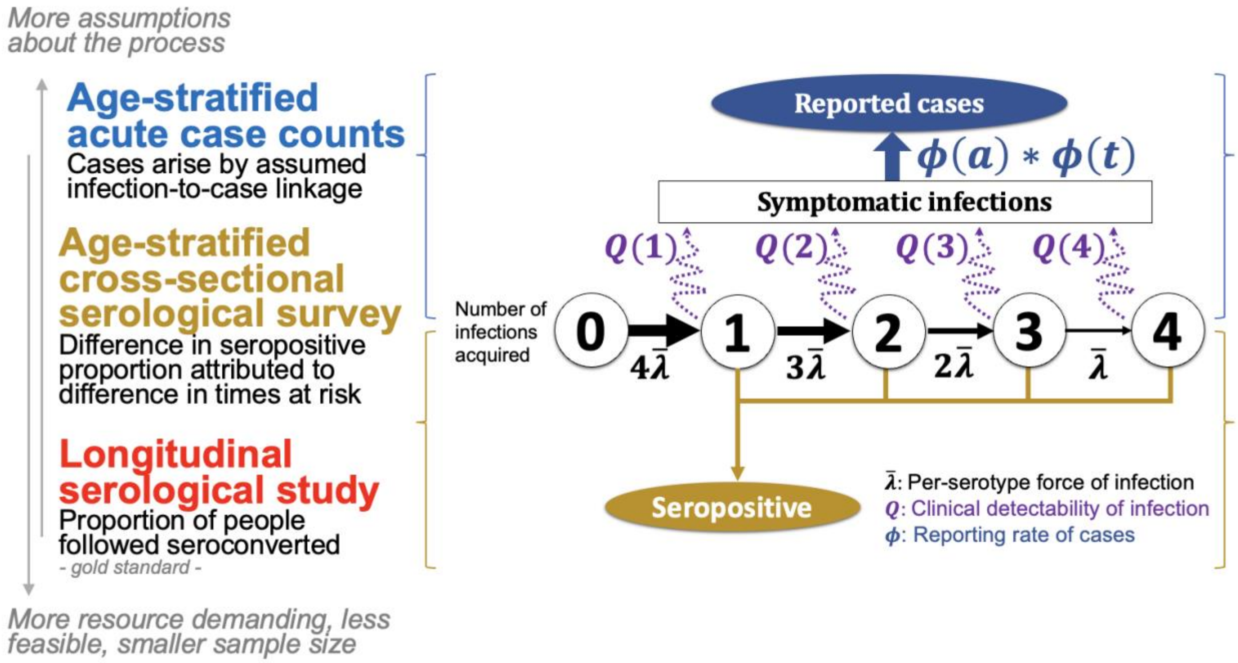

**Figure S1. Diagram of standard models linking underlying infection process to observed data.**

**Table S2. Descriptions of parameters in the standard model linking force of infection to the number of dengue cases that sought care at Kamphaeng Phet Hospital (KPPH).**

| Parameter | Description | Prior |
|---|---|---|
| $\bar{\lambda}(t)$ | Annual per-serotype force of infection | Beta(2, 38) |
| $p_{severe}(1)$ | Probability that 1st infections of individuals resulted in severe infections relative to 2nd infections | Beta(1, 9) |
| $p_{severe}(i)$ | Probability that i-th infections of individuals resulted in severe infections relative to 2nd infections | Beta(1, 999) |
| $\phi(a)$ | Probability that a severe case of age a sought care at KPPH relative to cases of age 0-2yrs (reference class) | Lognormal(0, 0.1) |
| $\phi(t)$ | Probability that a severe case at age 0-2yrs sought care at KPPH at time t | Beta(2,2) |

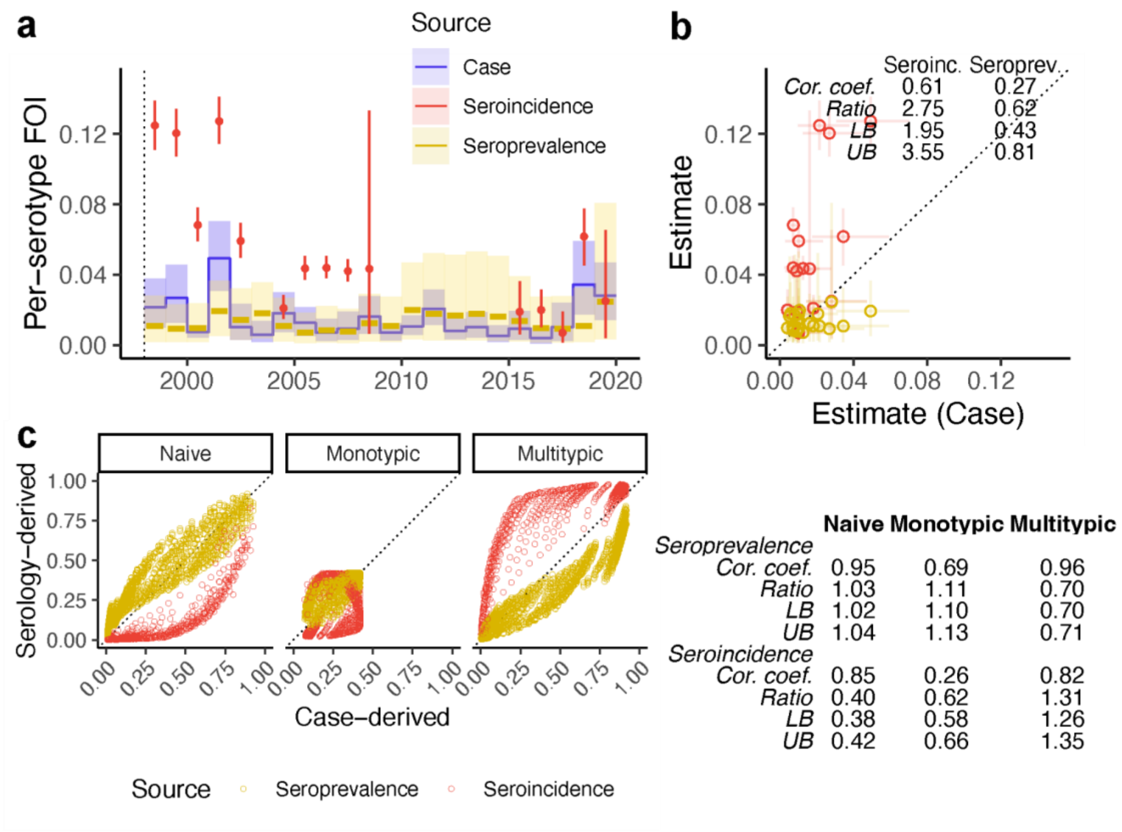

**Figure S2. Estimates from standard force of infection (FOI) inference models when using seropositivity threshold of 20. a)** Annual FOI estimated from each of the data sources: seroincidence data (red) and seroprevalence data (yellow) using seropositivity threshold of GMT>=20, and case data. **b)** Seroincidence-derived (red) and seroprevalence-derived FOI (yellow) compared against case-derived FOI (x-axis) and **c)** relationships between the respective susceptibility reconstructions. Each point in the reconstruction represents the proportion in each age-year that has not been infected with DENV (naive), has been infected by one serotype (monotypic) or more than one serotype (multitypic). LB=Lower bound, UB=Upper bound of the 95%CI of ratios.

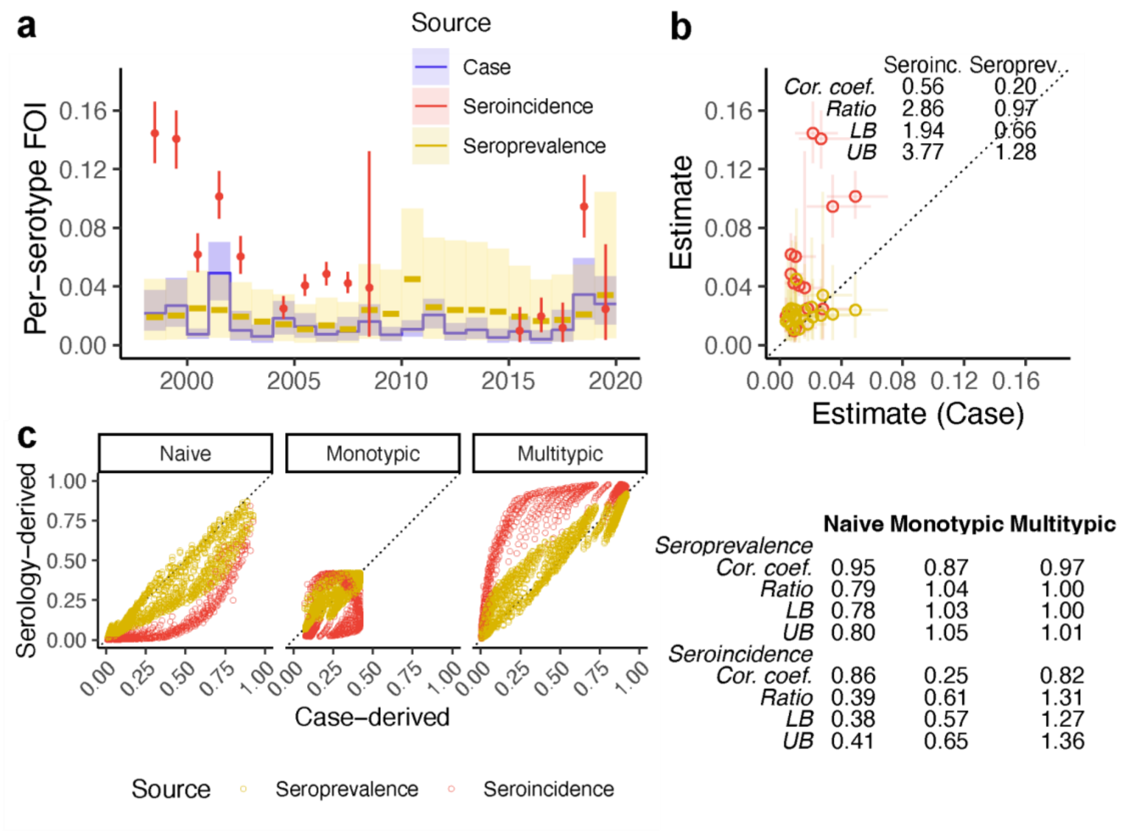

**Figure S3. Case-derived force of infection (FOI) inferred from lab-confirmed cases** compared against seroprevalence and seroincidence derived FOIs when using seropositivity threshold of 10. **a)** Annual FOI estimated from each of the data sources: seroincidence data (red) and seroprevalence data (yellow) using seropositivity threshold of GMT>=10, and lab-confirmed case data. **b)** Seroincidence-derived (red) and seroprevalence-derived FOI (yellow) compared against case-derived FOI (x-axis) and **c)** relationships between the respective susceptibility reconstructions. Each point in the reconstruction represents the proportion in each age-year that has not been infected with DENV (naive), has been infected by one serotype (monotypic) or more than one serotype (multitypic). LB=Lower bound, UB=Upper bound of the 95%CI of ratios.

### Simulations to study effects of model assumption violations

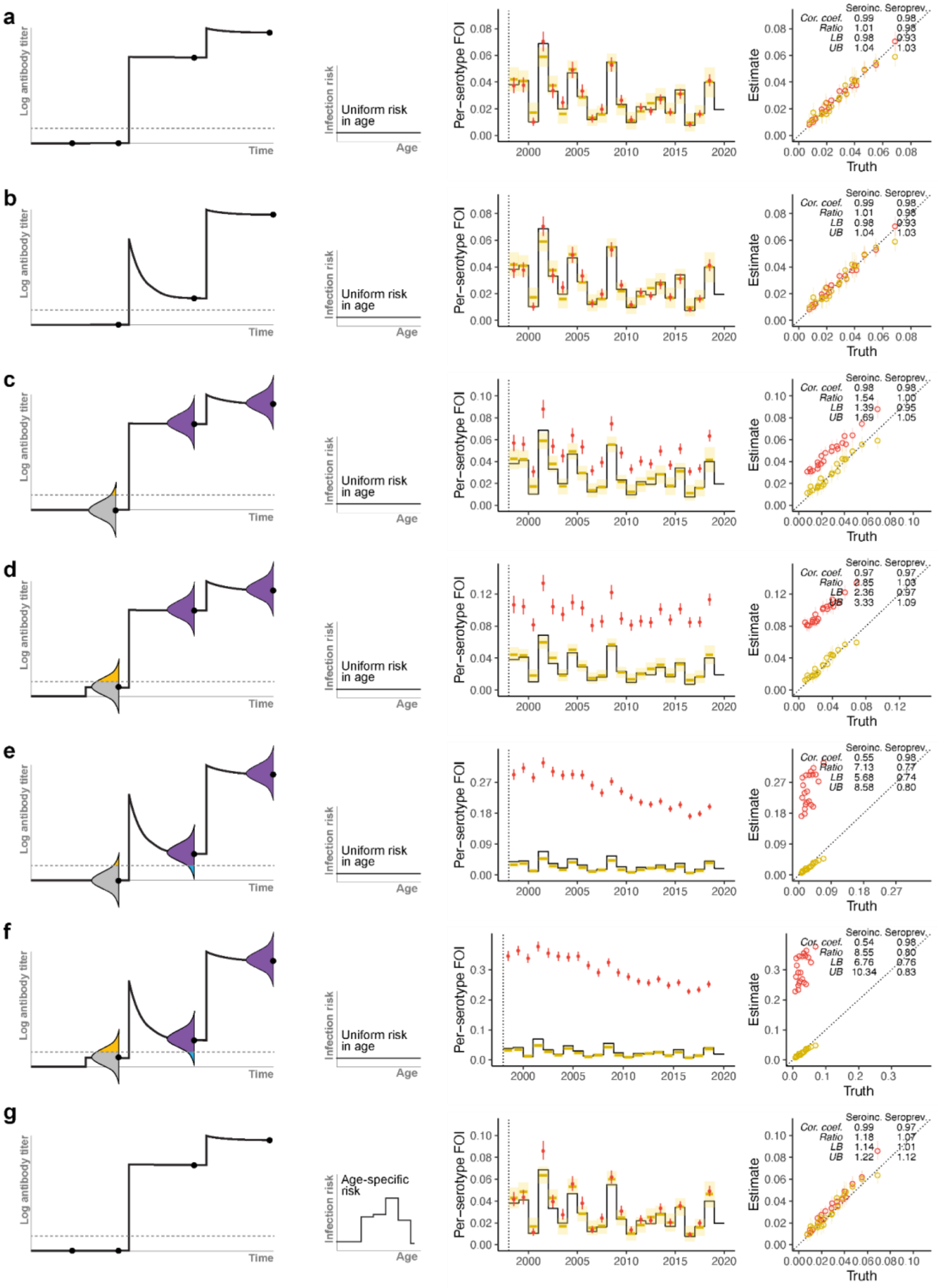

**Figure S4. Effects of violating model assumptions on inferred infection risk in highly powered datasets using standard serological models at a low seropositivity threshold (GMT>=10).** Left of each panel are schematics of assay variability, antibody kinetics, and seropositivity thresholds used to simulate the data: **a)** Assay without noise, durable monotypic titers, without cross-reactive (CXR) titers, **b)** assay without noise, waning monotypic titers, without CXR titers, **c)** noisy assay, durable monotypic titers, without CXR titers, **d)** noisy assay, durable monotypic titers, with CXR titers, **e)** noisy assay, waning monotypic titers, without CXR titers, **f)** noisy assay, waning monotypic titers, with CXR titers. All of which infection risk is uniform in age. **g)** Assay without noise, durable monotypic titers, without CXR titers, but infection risk is non-uniform in age. Center of each panel compares inferred force of infection from seroincidence data (red) and seroprevalence data (yellow) to ground truth (black). Right of the panels are scatter plots between inferred infection risk and true infection risk by age.

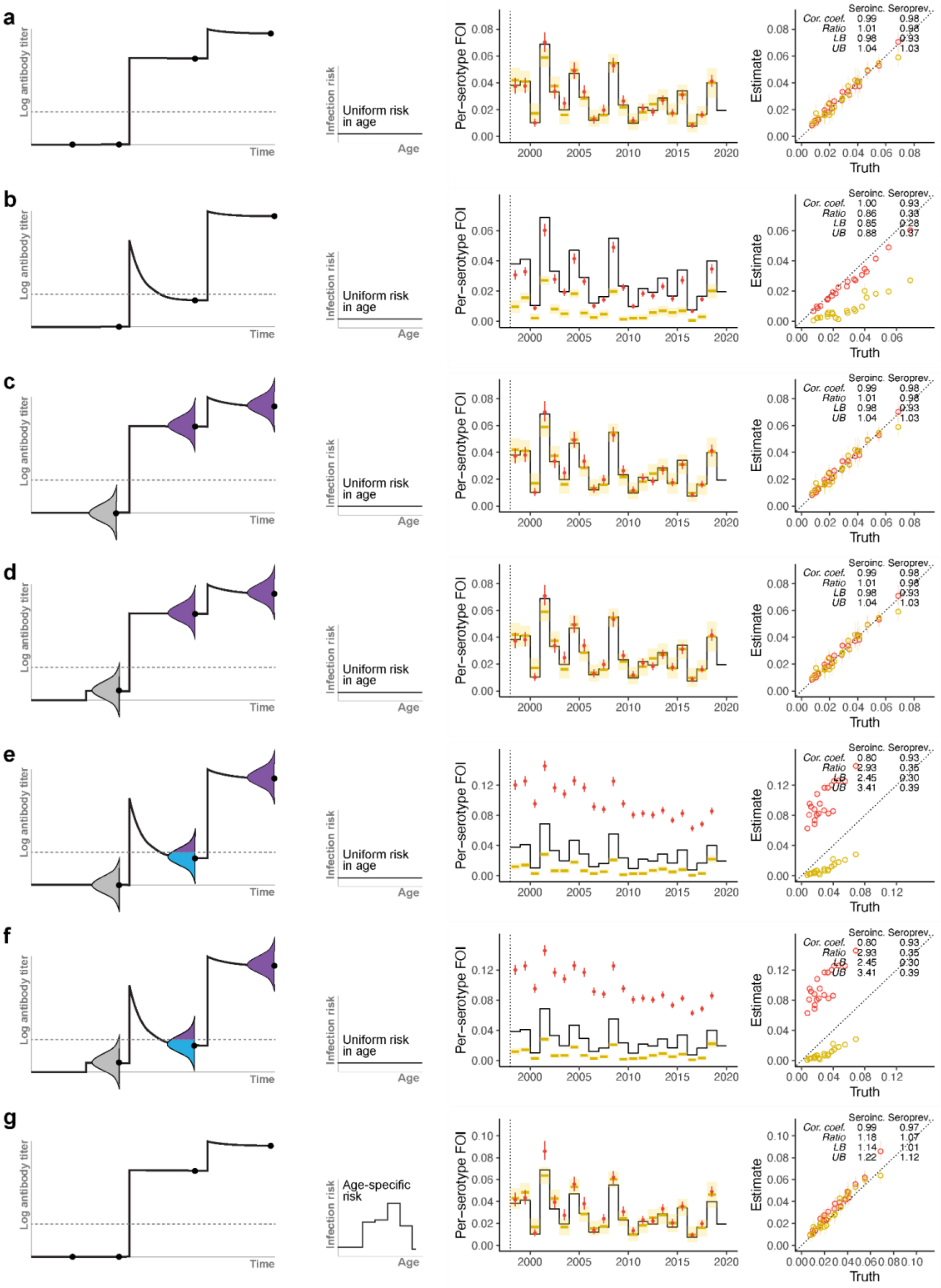

**Figure S5. Effects of violating model assumptions on inferred infection risk in highly powered datasets using standard serological models at a high seropositivity threshold (GMT>=20).** Left of each panel are schematics of assay variability, antibody kinetics, and seropositivity thresholds used to simulate the data: **a)** Assay without noise, durable monotypic titers, without cross-reactive (CXR) titers, **b)** assay without noise, waning monotypic titers, without CXR titers, **c)** noisy assay, durable monotypic titers, without CXR titers, **d)** noisy assay, durable monotypic titers, with CXR titers, **e)** noisy assay, waning monotypic titers, without CXR titers, **f)** noisy assay, waning monotypic titers, with CXR titers. All of which infection risk is uniform in age. **g)** Assay without noise, durable monotypic titers, without CXR titers, but infection risk is non-uniform in age. Center of each panel compares inferred force of infection from seroincidence data (red) and seroprevalence data (yellow) to ground truth (black). Right of the panels are scatter plots between inferred infection risk and true infection risk by age.

#### Efficiency in correcting for model violations

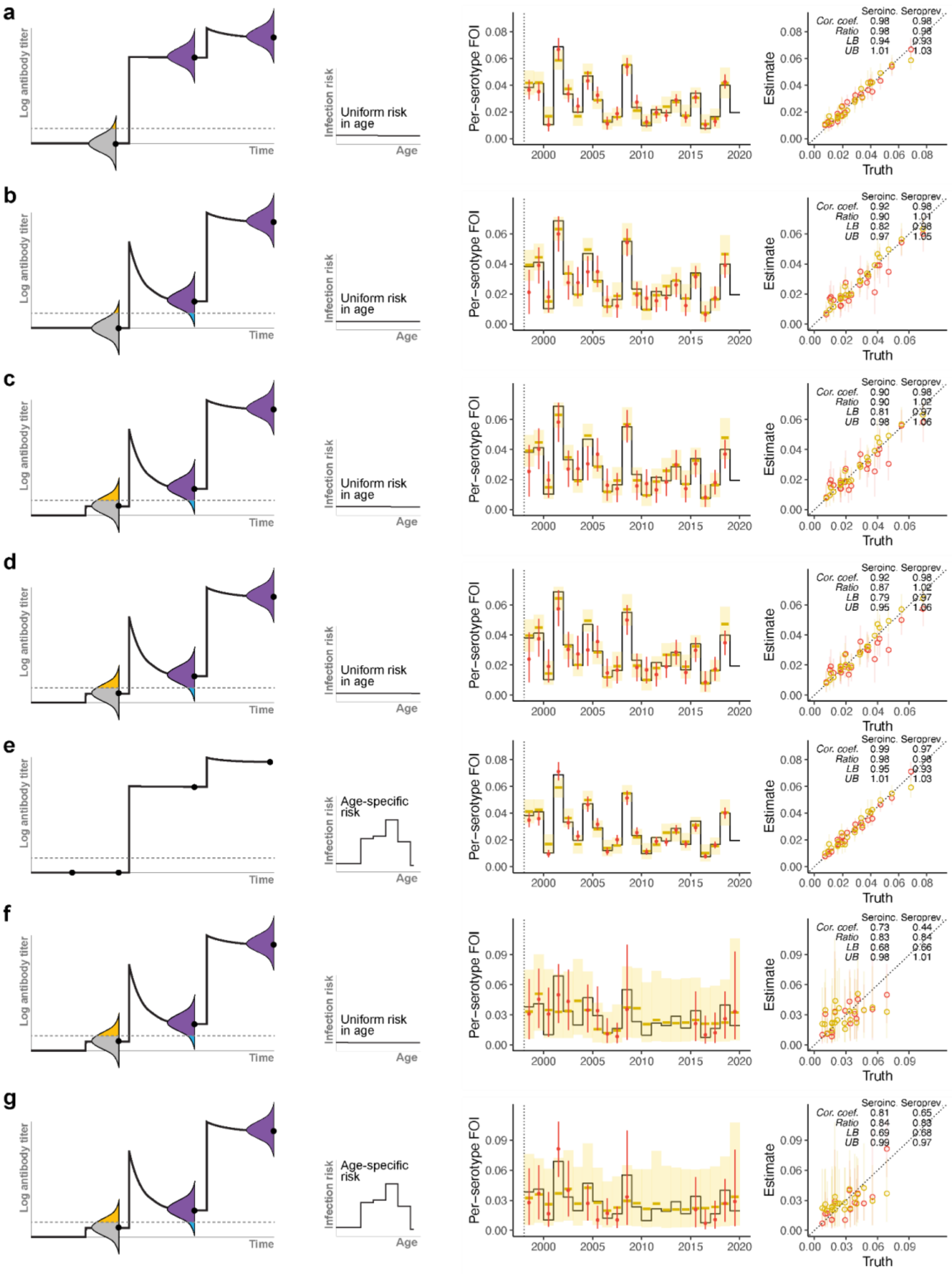

**Figure S6. Efficiency in correcting for model assumption violations to recover true temporal infection risk when using a low seropositivity threshold (GMT>=10).** Left of each panel are schematics of assay variability, antibody kinetics, seropositivity thresholds, and age-specific infection risk used to simulate the data: **a)** noisy assay, durable monotypic titers, without cross-reactive titers, uniform risk in age **b)** noisy assay, waning monotypic titers, without cross-reactive titers, uniform risk in age **c)** noisy assay, waning monotypic titers, with cross-reactive titers, uniform risk in age **d)** noisy assay, waning monotypic titers, with cross-reactive titers, non-uniform risk in age, **e)** assay without noise, durable monotypic titers, without cross-reactive titers, non-uniform risk in age. All of which were simulated as highly powered datasets. **f,g)** Analogs of (c,d) but simulated with power matched that of the cohort studies in Kamphaeng Phet. Center of each panel compares inferred temporal force of infection from seroincidence data (red) and seroprevalence data (yellow) to ground truth (black). Right of the panels are scatter plots between inferred infection risk and true infection risk by age.

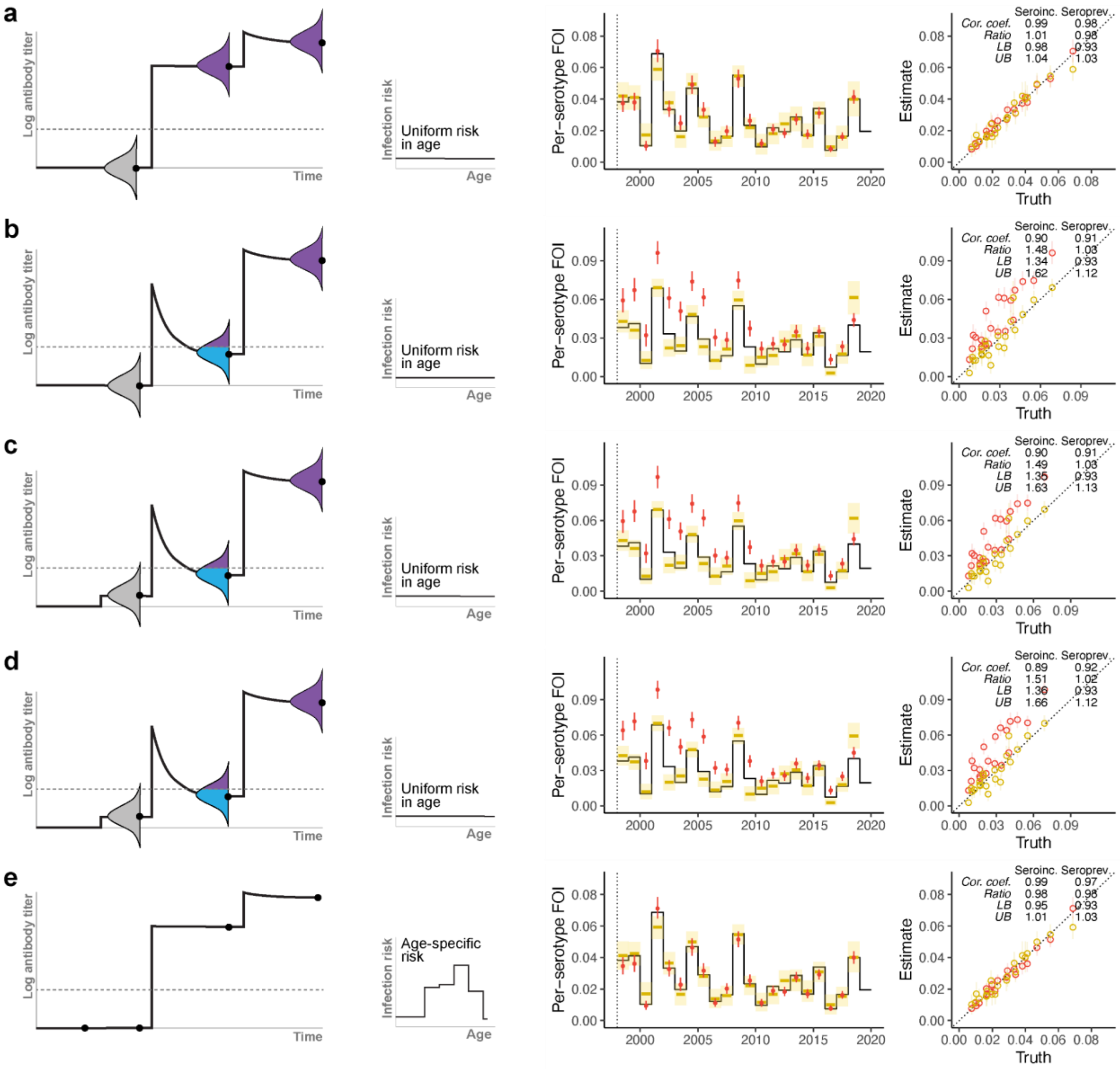

**Figure S7. Efficiency in correcting for model assumption violations to recover true temporal infection risk when using a high seropositivity threshold (GMT>=20).** Left of each panel are schematics of assay variability, antibody kinetics, seropositivity thresholds, and age-specific infection risk used to simulate the data: **a)** noisy assay, durable monotypic titers, without cross-reactive titers, uniform risk in age **b)** noisy assay, waning monotypic titers, without cross-reactive titers, uniform risk in age **c)** noisy assay, waning monotypic titers, with cross-reactive titers, uniform risk in age **d)** noisy assay, waning monotypic titers, with cross-reactive titers, non-uniform risk in age, **e)** assay without noise, durable monotypic titers, without cross-reactive titers, non-uniform risk in age. All of which were simulated as highly powered datasets. Center of each panel compares inferred temporal force of infection from seroincidence data (red) and seroprevalence data (yellow) to ground truth (black). Right of the panels are scatter plots between inferred infection risk and true infection risk by age.

#### Extended models to reconcile infection risk estimates

**Table S3. Priors of parameters in the joint serology model.**

| Parameter | Description | Prior |
|---|---|---|
| $\tau(t)$ | Annual per-serotype force of infection faced by individuals in the reference age class | Beta(2, 38) |
| $\kappa(a)$ | Force of infection faced by a specific age class relative to individuals aged 0-5yrs (reference class) | Lognormal(0, 0.1) |
| $\Omega_{short,ref}$ | Short-term titer rise captured by post-interval bleeds of KPS1 for a 1st infection that occurred within the interval | Gamma(6.399141, 1.187225)* |
| $\Omega_{short,rel}(z)$ | Short-term titer rise captured by post-interval bleeds of study z for a 1st infection that occurred within the interval relative to KPS1 | Lognormal(0, 0.1) |
| $\Omega_{long}$ | Long-term titer rise after 1st infection of individuals | Gamma(0.9561622, 0.7189189)* |
| $\Omega_{0,rel}$ | Cross-reactive titer in DENV-naive individuals relative to $\Omega_{long}$ | Beta(1,9) |
| $\sigma$ | Standard deviation of titer measurements in sera of DENV-exposed individuals | Normal(0.49, 0.1)* |
| * Priors taken/solved from means and variances reported in Salje, 2018. | | |

**Table S4. Priors of parameters in the extended case-based model.**

| Parameter | Description | Prior |
|---|---|---|
| $\tau(t)$ | Annual force of infection faced by individuals in the reference age class | Beta(2, 38) |
| $\kappa(a)$ | Force of infection faced by a specific age class relative to individuals aged 0-2yrs (reference class) | Lognormal(0, 0.1) |
| $p_{severe}(1)$ | Probability that 1st infections of individuals resulted in severe infections relative to 2nd infections | Beta(1, 9) |
| $p_{severe}(i)$ | Probability that i-th infections (3rd or 4th) of individuals resulted in severe infections relative to 2nd infections | Beta(1, 999) |
| $\phi(a)$ | Probability that a severe case of age a sought care at KPPH relative to cases of age 0-2yrs (reference class) | Lognormal(0, 0.1) |
| $\phi(t)$ | Probability that a severe case of age 0-2yrs sought care at KPPH at year t | Beta(2,2) |

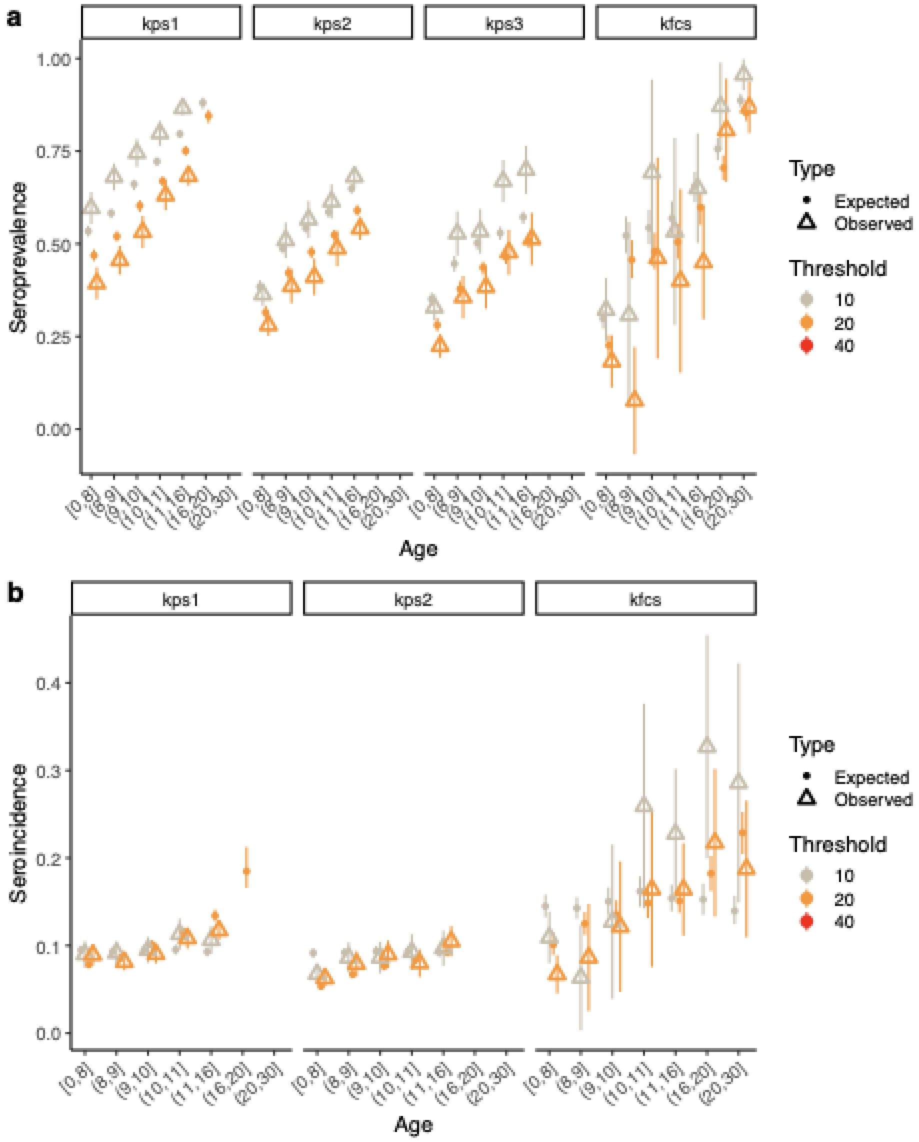

**Figure S8. Observed serology data vs expectations from the joint serology model fit. a)** Seropositive proportions by age group and study using seropositivity thresholds of GMT>=10 (gray) and 20 (orange) compared against their respective expectations from the model fits (lines). **b)** Seroincidence by age group and study using seropositivity threshold of 10.

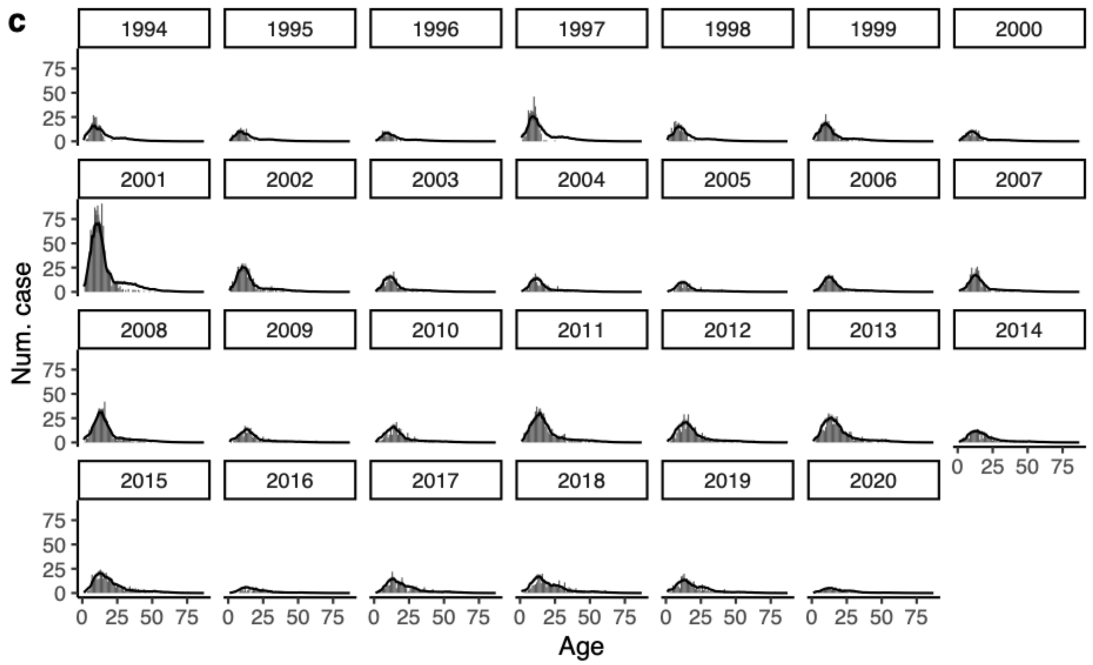

**Figure S9. Observed case data data vs expectations from the extended case-based model fit.** Number of reported dengue cases at KPPH by age and year (bars) compared against its expectations from the model fits (lines).

**Table S5. Posterior medians (and 95% credible intervals) of infection parameters in the joint serology model and extended case-based model.**

| Parameter and description | Year / Age group | Posterior median of joint serology model (95% credible interval) | Posterior median of case-based model (95% credible interval) |
|---|---|---|---|
| $\tau(t)$<br><br>Annual per-serotype force of infection faced by individuals in the reference age class | Up to 1984 | 0.041 (0.006, 0.127) | 0.020 (0.016, 0.023) |
| | 1985 | 0.034 (0.006, 0.102) | 0.168 (0.096, 0.240) |
| | 1986 | 0.022 (0.003, 0.065) | 0.108 (0.036, 0.196) |
| | 1987 | 0.064 (0.022, 0.109) | 0.053 (0.010, 0.121) |
| | 1988 | 0.040 (0.009, 0.083) | 0.025 (0.004, 0.072) |
| | 1989 | 0.066 (0.031, 0.101) | 0.029 (0.005, 0.080) |
| | 1990 | 0.027 (0.005, 0.057) | 0.028 (0.004, 0.083) |
| | 1991 | 0.024 (0.005, 0.051) | 0.026 (0.004, 0.072) |
| | 1992 | 0.027 (0.006, 0.055) | 0.019 (0.003, 0.059) |
| | 1993 | 0.041 (0.017, 0.064) | 0.022 (0.004, 0.065) |
| | 1994 | 0.010 (0.001, 0.027) | 0.036 (0.010, 0.070) |
| | 1995 | 0.013 (0.002, 0.033) | 0.022 (0.006, 0.042) |
| | 1996 | 0.028 (0.008, 0.051) | 0.014 (0.005, 0.026) |
| | 1997 | 0.017 (0.004, 0.036) | 0.042 (0.017, 0.077) |
| | 1998 | 0.032 (0.023, 0.041) | 0.014 (0.004, 0.030) |
| | 1999 | 0.027 (0.020, 0.034) | 0.018 (0.006, 0.037) |
| | 2000 | 0.001 (0.000, 0.004) | 0.006 (0.003, 0.010) |
| | 2001 | 0.031 (0.024, 0.038) | 0.037 (0.018, 0.061) |
| | 2002 | 0.002 (0.000, 0.006) | 0.009 (0.002, 0.021) |
| | 2003 | 0.032 (0.021, 0.043) | 0.005 (0.001, 0.012) |
| | 2004 | 0.003 (0.001, 0.008) | 0.008 (0.002, 0.018) |
| | 2005 | 0.007 (0.003, 0.011) | 0.005 (0.001, 0.013) |
| | 2006 | 0.015 (0.011, 0.020) | 0.007 (0.002, 0.015) |
| | 2007 | 0.007 (0.004, 0.011) | 0.009 (0.003, 0.019) |
| | 2008 | 0.015 (0.003, 0.034) | 0.017 (0.005, 0.033) |
| | 2009 | 0.012 (0.002, 0.031) | 0.009 (0.003, 0.017) |
| | 2010 | 0.036 (0.010, 0.064) | 0.025 (0.017, 0.035) |
| | 2011 | 0.018 (0.003, 0.050) | 0.042 (0.030, 0.058) |
| | 2012 | 0.018 (0.003, 0.051) | 0.007 (0.003, 0.014) |
| | 2013 | 0.022 (0.003, 0.059) | 0.008 (0.003, 0.016) |
| | 2014 | 0.018 (0.003, 0.051) | 0.005 (0.002, 0.010) |
| | 2015 | 0.005 (0.001, 0.013) | 0.007 (0.002, 0.015) |
| | 2016 | 0.003 (0.001, 0.009) | 0.005 (0.002, 0.012) |
| | 2017 | 0.002 (0.000, 0.007) | 0.007 (0.002, 0.015) |
| | 2018 | 0.042 (0.031, 0.053) | 0.021 (0.010, 0.036) |
| | 2019 | 0.016 (0.003, 0.045) | 0.018 (0.009, 0.031) |
| | 2020 | 0.042 (0.006, 0.137) | 0.008 (0.004, 0.013) |

| | | | |
|---|---|---|---|
| $\kappa(a)$<br><br>Force of infection faced by a specific age class relative to the reference class | 0-2 yrs<br>3-5 yrs<br>6-8 yrs<br>9-11 yrs<br>12-14 yrs<br>15-17 yrs<br>18-20 yrs<br>21-23 yrs<br>24-26 yrs<br>27-29 yrs<br>30-39 yrs<br>40-49 yrs<br>50-59 yrs<br>60-62 yrs<br>63+ yrs | Ref.<br>Ref.<br>1.189 (1.004, 1.388)<br>1.293 (1.127, 1.477)<br>1.099 (0.911, 1.318)<br>1.204 (1.000, 1.444)<br>1.000 (1.000, 1.000)<br>n/a<br>n/a<br>n/a<br>n/a<br>n/a<br>n/a<br>n/a<br>n/a | Ref.<br>1.070 (0.934, 1.220)<br>1.248 (1.091, 1.424)<br>1.276 (1.112, 1.451)<br>1.387 (1.215, 1.586)<br>1.214 (1.056, 1.389)<br>1.020 (0.888, 1.182)<br>0.910 (0.781, 1.062)<br>0.851 (0.727, 0.998)<br>0.865 (0.744, 1.015)<br>0.907 (0.769, 1.052)<br>0.741 (0.638, 0.857)<br>0.742 (0.644, 0.862)<br>0.919 (0.784, 1.068)<br>1.000 (1.000, 1.000) |

**Table S6. Posterior medians (and 95% credible intervals) of parameters in the joint serology model linking infection risk to serological data.**

| Parameter | Description | Posterior median (95% credible interval) |
|---|---|---|
| $\Omega_{short,ref}$ | Short-term titer rise captured by post-interval bleeds of KPS1 for a 1st infection that occurred within the interval | 7.823 (4.755, 12.894) |
| $\Omega_{short,rel}(z)$ | Short-term titer rise captured by post-interval bleeds of study z for a 1st infection that occurred within the interval relative to KPS1 | KPS2: 1.000 (0.823, 1.216)<br>KPS3: 0.999 (0.823, 1.220)<br>KFCS: 0.999 (0.817, 1.222) |
| $\Omega_{long}$ | Long-term titer rise after 1st infection of individuals | 2.728 (2.510, 2.941) |
| $\Omega_{0,rel}$ | Cross-reactive titers in DENV-naive individuals relative to long-term titer rise after 1st infection of individuals | 0.097 (0.016, 0.193) |
| $\sigma$ | Standard deviation of titer measurements in sera of DENV-exposed individuals | 0.505 (0.354, 0.647) |

**Table S7. Posterior medians (and 95% credible intervals) of parameters in the case-based model linking infection risk to case data.**

| Parameter | Description | Posterior median (95% credible interval) | |
|---|---|---|---|
| $p_{severe}(i)$ | Probability that i-th infections of individuals resulted in severe infections relative to 2nd infections | i=1 | 0.054 (0.028, 0.081) |
| | | i=2 | Ref. |
| | | i=3 | 0.001 (0.000, 0.004) |
| | | i=4 | 0.001 (0.000, 0.005) |
| $\phi(a)$ | Probability that a severe case of age a sought care at KPPH relative to cases of age 0-2yrs (reference class) | 0-2 | Ref. |
| | | 3-5 | 1.122 (0.965, 1.301) |
| | | 6-8 | 1.202 (1.036, 1.394) |
| | | 9-11 | 1.294 (1.116, 1.486) |
| | | 12-14 | 1.123 (0.972, 1.284) |
| | | 15-17 | 1.017 (0.883, 1.184) |
| | | 18-20 | 0.875 (0.758, 1.013) |
| | | 21-23 | 0.712 (0.615, 0.826) |
| | | 24-26 | 0.752 (0.641, 0.872) |
| | | 27-29 | 0.801 (0.679, 0.936) |
| | | 30-39 | 0.800 (0.688, 0.933) |
| | | 40-49 | 0.915 (0.779, 1.071) |
| | | 50-59 | 1.001 (0.850, 1.172) |
| | | 60+ | 1.401 (1.166, 1.670) |
| $\phi(t)$ | Probability that a severe case at age 0-2yrs sought care at KPPH at year t | 1994-1995 | 0.020 (0.010, 0.071) |
| | | 1996-1997 | 0.025 (0.013, 0.065) |
| | | 1998-1999 | 0.041 (0.020, 0.139) |
| | | 2000-2001 | 0.073 (0.044, 0.149) |
| | | 2002-2003 | 0.116 (0.047, 0.456) |
| | | 2004-2005 | 0.071 (0.030, 0.284) |
| | | 2006-2007 | 0.082 (0.039, 0.284) |
| | | 2008-2009 | 0.097 (0.049, 0.307) |
| | | 2010-2011 | 0.046 (0.033, 0.066) |
| | | 2012-2013 | 0.252 (0.129, 0.633) |
| | | 2014-2015 | 0.200 (0.096, 0.588) |
| | | 2016-2017 | 0.181 (0.084, 0.554) |
| | | 2018-2020 | 0.058 (0.034, 0.122) |

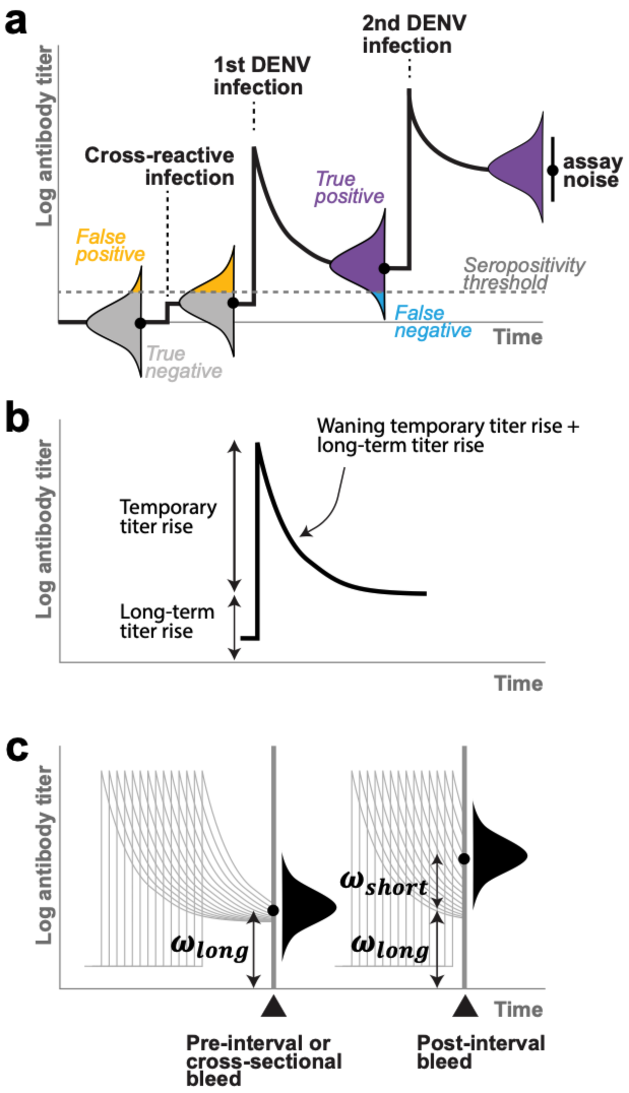

**Figure S10. Relationship between titer parameters in the joint serology model and individual-level anti-DENV antibody kinetics** described Figure 2 and in the Supplementary Mathematical Analysis. **a)** Illustration of anti-DENV antibody kinetics as an individual acquires a cross-reactive (CXR) virus infection or vaccination (i.e., not DENV), one DENV infection, and >1 DENV infections. Measured titers distribute around the true underlying titers with variability depending on the assay characteristics. **b)** Titer components in the antibody kinetics upon first DENV infection of an individual. **c)** Titer parameters in the joint serology model encodes the average long-term titer rises captured in pre-interval or cross-sectional blood samples ($\Omega_{long}$) across individuals, and the average titer rises captured in post-interval blood samples as a result of infections that occurred during the interval ($\Omega_{long} + \Omega_{short}$).

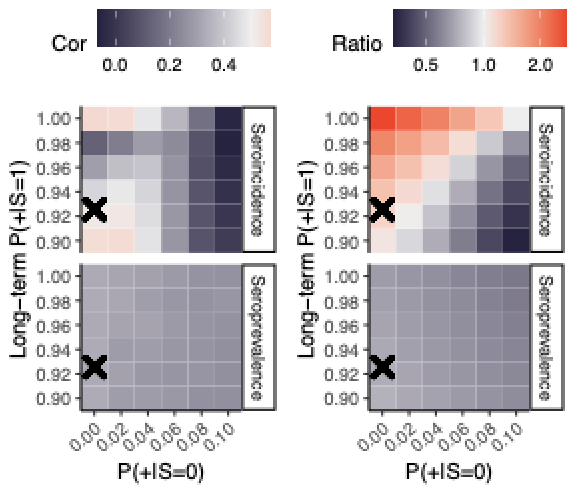

**Figure S11. Variations in FOI congruence across presumed test positive probabilities.** Effects of presumed test positivity probabilities on the correlation and ratio between temporal FOIs inferred from the extended case-based model and temporal FOIs inferred from a single data source (either seroincidence or seroprevalence at seropositivity threshold of 20) imposed with age-specific risk inferred from the extended case-based model. Test positive probabilities estimated from the joint serology model are annotated as crosses for comparison.
