## Supplementary Mathematical Analysis for "Reconciling heterogeneous dengue virus infection risk estimates from different study designs"

### Supplementary mathematical analysis: biases in force of infection inferred from seroincidence data

Notations used throughout this document are defined as follows.

$$\begin{aligned}
P(\ominus_t) &= \text{Probability of testing negative at time } t\\
P(\oplus_t) &= \text{Probability of testing positive at time } t\\
P(\ominus_t^T) &= \text{Probability of being true negative at time } t\\
P(\oplus_t^T) &= \text{Probability of being true positive at time } t\\
P(\ominus_t^F) &= \text{Probability of being false negative at time } t\\
P(\oplus_t^F) &= \text{Probability of being false positive at time } t\\
P(S_t = i) &= \text{Probability of having acquired } i \text{ infections at time } t\\
p_a &= \text{Probability of escaping a particular serotype up till age } a\\
\bar{\lambda} &= \text{Average historical per-serotype force of infection}\\
\lambda_t &= \text{Per-serotype force of infection during time interval } (t, t+\Delta t]\\
\omega &= \text{Long-term persistent titer rise acquired upon primary dengue infection}\\
\gamma &= \text{Temporary titer rise upon primary dengue infection}\\
\delta &= \text{Rate of exponential decay in temporary titers}\\
\sigma &= \text{Standard deviation of assay measurements from true underlying titer of a DENV-exposed individual}\\
\sigma_0 &= \text{Standard deviation of assay measurements from true underlying titer of a DENV-naive individual}\\
\nu &= \text{Seropositivity titer cut-off}\\
\Phi(x) &= \text{Cumulative density of a standard normal distribution at value } x
\end{aligned}$$

Longitudinal serology is typically viewed as the gold standard for measuring levels of transmission within a time period as it directly measures serological changes of individuals. We can express the **true probability of first infection incidence** during a time interval $(t, t + \Delta t]$ as

$$P(S_{t+\Delta t} \geq 1 | S_t = 0) = P(S_{t+\Delta t} = 1 | S_t = 0) + P(S_{t+\Delta t} > 1 | S_t = 0) \tag{1}$$

**Probability of observing seroconversion** in an individual, $P(\oplus_{(t,t+\Delta t]} | \ominus_t)$, can be expanded to a weighted average between probabilities of seroconverting given being truely negative at the first time point, $t$, and being falsely negative.

$$P(\oplus_{(t,t+\Delta t]} | \ominus_t) = \frac{P(\ominus_t^T)}{P(\ominus_t)}[P(\oplus_{t+\Delta t}^T | \ominus_t^T) + P(\oplus_{t+\Delta t}^F | \ominus_t^T)] + \frac{P(\ominus_t^F)}{P(\ominus_t)}[P(\oplus_{t+\Delta t}^T | \ominus_t^F) + P(\oplus_{t+\Delta t}^F | \ominus_t^F)] \tag{2}$$

**Biases in seroconversion probabilities when truly negative at t** from the true probability of first infection incidence is then

$$
\begin{aligned}
&[P(\oplus^T_{(t,t+\Delta t]}|\ominus^T_t) + P(\oplus^F_{(t,t+\Delta t]}|\ominus^T_t)] - P(S_{(t,t+\Delta t]} = 1|S_t = 0) - P(S_{(t,t+\Delta t]} > 1|S_t = 0) \\
&= P(S_{(t,t+\Delta t]} = 1|S_t = 0)\ P(\oplus^T_{(t,t+\Delta t]}|S_{(t,t+\Delta t]} = 1) + P(S_{(t,t+\Delta t]} > 1|S_t = 0)\ P(\oplus^T_{(t,t+\Delta t]}|S_{(t,t+\Delta t]} > 1) \\
&+ P(S_{(t,t+\Delta t]} = 0|S_t = 0)\ P(\oplus^F_{(t,t+\Delta t]}|S_{(t,t+\Delta t]} = 0) - P(S_{(t,t+\Delta t]} = 1|S_t = 0) - P(S_{(t,t+\Delta t]} > 1|S_t = 0) \\
&= P(S_{(t,t+\Delta t]} = 0|S_t = 0)\ P(\oplus^F_{(t,t+\Delta t]}|S_{(t,t+\Delta t]} = 0) \\
&+ P(S_{(t,t+\Delta t]} = 1|S_t = 0)[P(\oplus^T_{(t,t+\Delta t]}|S_{(t,t+\Delta t]} = 1) - 1] \\
&+ P(S_{(t,t+\Delta t]} > 1|S_t = 0)[P(\oplus^T_{(t,t+\Delta t]}|S_{(t,t+\Delta t]} > 1) - 1]
\end{aligned} \tag{3}
$$

We can see that the bias is a weighted average between biases when no infection occurred during the interval ($S_{(t,t+\Delta t]} = 0, S_t = 0$), when one infection occurred during the interval ($S_{(t,t+\Delta t]} = 1|S_t = 0$), and when more than one infection occurred during the interval ($S_{(t,t+\Delta t]} > 1|S_t = 0$). Assuming long-lived protection against infecting serotypes and no cross-protection between the serotypes, we can write the probabilities of these scenarios (i.e., the weights) for an individual of age $a$ at time $(t, t + \Delta t]$ as

$$
P(S_{(t,t+\Delta t]} = i) = \binom{4}{i}\ (e^{-\bar{\lambda} a})^{4-i}\ (1 - e^{-\bar{\lambda} a})^i \tag{4}
$$

Previous studies on the kinetics of anti-DENV antibodies have shown that the rise in antibody levels after having acquired one infection, i.e., monotypic titers, consists of a portion which diminishes within a year and a portion which persists for longer (cite: Leah, Henrik). Following Salje et al, the kinetics of $H_t$, the true titer at time $t$, can be characterized as

$$
H_t = \omega + \gamma e^{-(t-\star t)\delta} \tag{5}
$$

where $\omega$ is the long-term persistent titer acquired upon infection, $\gamma$ is the temporary titer rise, $\delta$ is the rate of exponential decay in temporary titer rise, and $\star t$ is time at which the infection occurred. For an assay with measurement noise which follows a normal distribution $N(0, \sigma^2)$, using seropositivity cut-off $\nu$, the probability of testing positive after being infected once is

$$
P(\oplus^T_t|S_t = 1) = 1 - \Phi(\frac{\nu - H_t}{\sigma}) \tag{6}
$$

In DENV-naive individuals, although anti-DENV antibodies are absent, cross-reactive titers against other *Flaviviruses* in circulation could plausibly result in non-zero titers against DENV, $\omega_0$. We can, in a similar manner, express the probability of testing positive when DENV-naive as

$$
P(\oplus^F_t|S_t = 0) = 1 - \Phi(\frac{\nu - \omega_0}{\sigma}) \tag{7}
$$

Let probability of escaping a particular serotype during time interval $e^{-\lambda_t \Delta t} = p$, we can express (3) as

$$
\begin{aligned}
&p^4\ [1 - \Phi(\frac{\nu - \omega_0}{\sigma})] + \binom{4}{1}\ p^3\ (1-p)\ [(1 - \Phi(\frac{\nu - H_t}{\sigma})) - 1] + [1 - p^4 - \binom{4}{1}\ p^3\ (1-p)]\ [1 - 1] \\
&= p^4\ [1 - \Phi(\frac{\nu - \omega_0}{\sigma})] - 4\ p^3\ (1-p)\ \Phi(\frac{\nu - H_t}{\sigma}))
\end{aligned} \tag{8}
$$

For the first case in Equation (3) where no infection occurred, the bias increases when the seropostivity threshold is lowered. In the second case where only one infection occurred, the bias becomes less negative when time since infection is small or when the seropostivity threshold is lowered. Since time since infection in this case is bounded by the interval length, shorter bleeding intervals are more likely to be less negative. Titers in individuals who have acquired multiple DENV infections, i.e., multitypic titers, are more robust and can be assumed to test negative at negligible levels. It follows that minimal bias would arise from the last case where more than one infection occurred during the interval.

**Biases in seroconversion probabilities when falsely negative at t**, under these described processes, can be expressed as

$$\begin{aligned} &P(\oplus^T_{(t,t+\Delta t]}|\ominus^F_t) + P(\oplus^F_{(t,t+\Delta t]}|\ominus^F_t) - P(S_{(t,t+\Delta t]} = 1|S_t = 0) - P(S_{(t,t+\Delta t]} > 1|S_t = 0) \\ &= P(\oplus^T_{(t,t+\Delta t]}|\ominus_t, S_t = 1) + 0 - P(S_{(t,t+\Delta t]} = 1|S_t = 0) - P(S_{(t,t+\Delta t]} > 1|S_t = 0) \\ &= P(\oplus_{(t,t+\Delta t]}, S_{(t,t+\Delta t]} = 1|\ominus_t, S_t = 1) + P(\oplus^T_{(t,t+\Delta t]}, S_{(t,t+\Delta t]} > 1|\ominus^F_t, S_t = 1) \\ &- P(S_{(t,t+\Delta t]} = 1|S_t = 0) - P(S_{(t,t+\Delta t]} > 1|S_t = 0) \end{aligned} \tag{9}$$

The first case where $S_{(t,t+\Delta t]} = S_t = 1$ means that no infection occurred during the interval. $P(\oplus_{(t,t+\Delta t]})$ in this case has a lowerbound of $1 - \Phi(\frac{\nu-\omega}{\sigma})$. In the second case where infection(s) occurred during the interval, $S_t = 1$ and $S_{(t,t+\Delta t]} > 1$, $P(\oplus_{(t,t+\Delta t]}) \approx 1$. Hence, $P(\oplus^T_{(t,t+\Delta t]}|\ominus^F_t)$ is a weighted average between two quantities, $1 - \Phi(\frac{\nu-\omega}{\sigma})$ and 1. Longer time between blood draws, $\Delta t$, and higher FOI, $\lambda_t$, during the interval tends the quantity towards 1. Again, for probability of escaping a particular serotype during the interval $p = e^{-\lambda_t \Delta t}$, we can express (9) as

$$\begin{aligned} &p^3 * (1 - \Phi(\frac{\nu - H_t}{\sigma})) + (1 - p^3) - (1 - p^4) \\ &= p^4 - \Phi(\frac{\nu - H_t}{\sigma})p^3 \end{aligned} \tag{10}$$

**Relative contributions of the biases** can be expressed as

$$\begin{aligned} \frac{P(\ominus^F_t)}{P(\ominus^T_t)} &= \frac{P(S_t = 1)\ P(\ominus^F_t|S_t = 1)}{P(S_t = 0)\ P(\ominus^T_t|S_t = 0)} \\ &= \frac{\binom{4}{1}\ (e^{-\bar{\lambda}a})^3\ (1 - e^{-\bar{\lambda}a})}{(e^{-\bar{\lambda}a})^4}\ \frac{P(\ominus^F_t|S_t = 1)}{P(\ominus^T_t|S_t = 0)} \\ &= 4\ \frac{(1 - e^{-\bar{\lambda}a})}{(e^{-\bar{\lambda}a})}\ \frac{P(\ominus^F_t|S_t = 1)}{P(\ominus^T_t|S_t = 0)} \end{aligned} \tag{11}$$

The quantity is a product of the constant 4 and two variable components. The first variable component is zero when $a$ is zero or the average per-serotype force of infection $\bar{\lambda}$ is zero and grows as the product $\bar{\lambda}a$ increases. The second variable component depends on the recency of the dengue infection with bounds

$$\frac{\Phi(\frac{\nu-\omega-\gamma}{\sigma})}{\Phi(\frac{\nu-\omega_0}{\sigma})} \leq \frac{P(\ominus^F_t|S_t = 1)}{P(\ominus^T_t|S_t = 0)} \leq \frac{\Phi(\frac{\nu-\omega}{\sigma})}{\Phi(\frac{\nu-\omega_0}{\sigma})} \leq 1 \tag{12}$$

Assuming cross-reactive titer $\omega_0$ is at most $\omega$, the quantity is bounded at one. Intuitively, lowering the positivity cut-off $\nu$ would decrease contribution of $\ominus^F_t$ to $\ominus_t$. However, the contribution can only be minimized down to $1/(1 + \Phi(\frac{\nu-\omega_0}{\sigma})/\Phi(\frac{\nu-\omega-\gamma}{\sigma}))$ and the efficiency of such reduction declines with age and historical FOI $\bar{\lambda}$.

Figure S1 illustrates ranges of the biases and their relative contributions using mean parameter estimates of $\omega = 1.33$ and $\sigma = 0.49$ from Salje, 2018 under various interval lengths and seropositivity cut-offs. The amount of cross-reactive titers relative to DENV-specific titers was artibrarily set to 20% ($\omega_0 = 0.2\omega$). Per-serotype FOI (historical and during the interval) is set to 0.03. Under these parameters, we can see that the magnitude

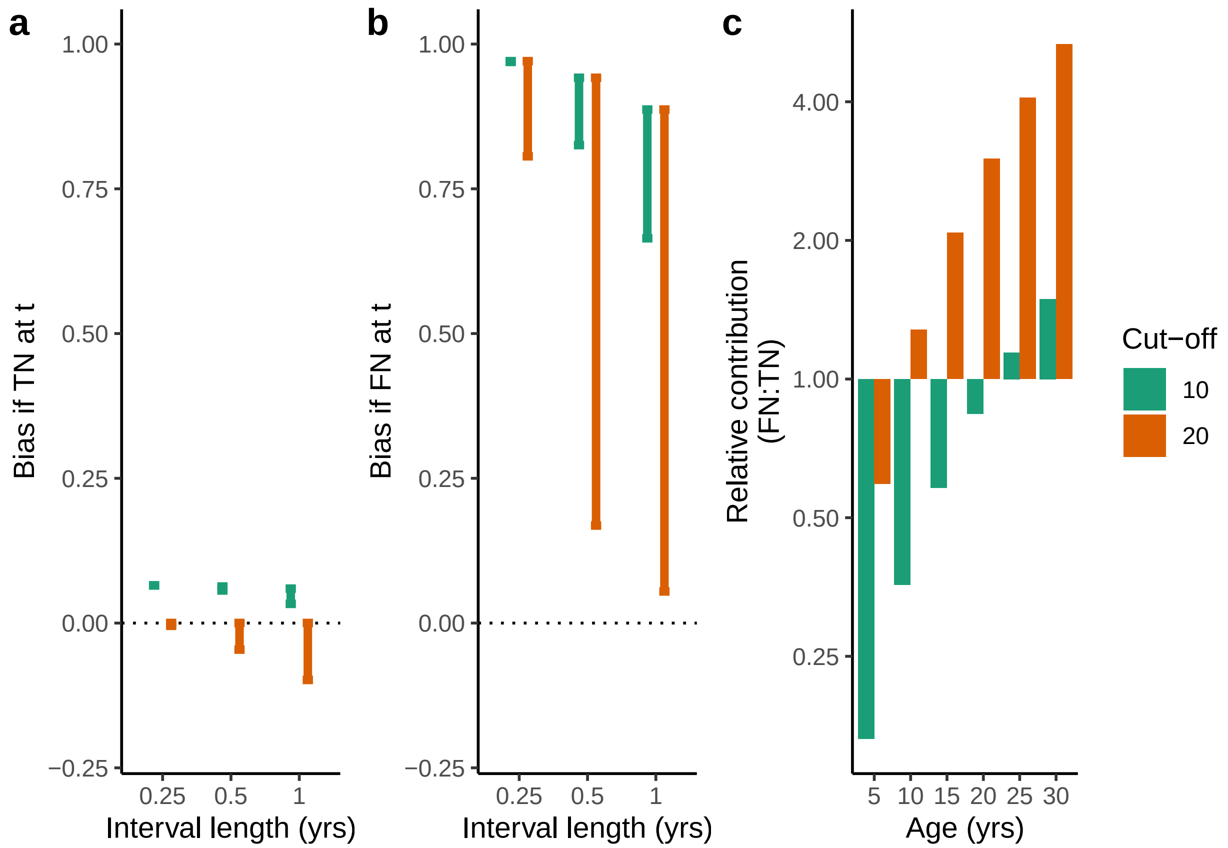


Figure 1: Biases in seroconversion probabilities illustrated using antibody kinetics estimates from Salje, 2018. Biases in the case where the individual was a) true negative (TN) at pre-interval bleed versus b) false negative (FN) at pre-interval bleed. c) Upper bound of relative contribution between the FN and TN case to the seroconversion probability as a function of pre-interval age for seropositivity cut-offs 10 and 20.

of positive bias from false negative scenario at $t$ is far higher biases from the true negative scenario. Increasing the cut-off reduces the lower bound of such inflation (leading to greater uncertainty in the amount of bias introduced) and increases the contribution of the false negative scenario.

**Effects of seasonality on test positivity.** The derivations, so far, assumes that FOI is constant throughout the year. However, it is conceivable that timings of the bleeds in relation to phases in the dengue seasons would lead to non-uniform distribution of the infections, and hence, may influence the expected biases.

Let the per-serotype FOI at age $a$ of an individual $\lambda(a) = \bar{\lambda} + \delta_\lambda cos(2\pi(a + \delta_t))$ where $\delta_\lambda$ is the magnitude of fluctuation and $\delta_t$ governs the phase of the fluctuation in relation to age. The per-serotype infection risk faced up till age $A$ is then

$$\Lambda(A) = \int_0^A \bar{\lambda} + \delta_\lambda cos(2\pi(a + \delta_t))\, da = \bar{\lambda} A + \frac{\delta_\lambda}{2\pi}[sin(2\pi(A + \delta_t)) - sin(2\pi\delta_t)] \tag{13}$$

We can write the probability that the individual was infected by a particular serotype at age $A$ as $e^{-\Lambda(A)}\lambda(A)$. Thus, the expected underlying titer of the individual at age $A_1$ given the individual was monotypically infected at that point is

$$\begin{aligned} \mathbb{E}[H_{A_1}|S_{A_1} = 1] &= \frac{3\ e^{-\Lambda(A_1)}\ \int_0^{A_1}(\omega + \gamma e^{-\delta(A_1 - A)})\ e^{-\Lambda(A)}\lambda(A)\, dA}{3\ e^{-\Lambda(A_1)}\ (1 - e^{-\Lambda(A_1)})} \\ &= \frac{\int_0^{A_1}(\omega + \gamma e^{-\delta(A_1 - A)})\ e^{-\Lambda(A)}\lambda(A)\, dA}{1 - e^{-\Lambda(A_1)}} \end{aligned} \tag{14}$$

The growing contribution of $\bar{\lambda}A$ in $\Lambda(A)$ as age increases shrinks the impact of timing of the pre-interval blood draw in relation to seasonality. Figure S2a-c illustrates this shrinkage under parameter estimates from Salje, 2018. On the other hand, for infections that occurred within the intervals, seasonality and the interval lengths dictates the distribution of time since infection at the second bleed (Figure S2d). These timing variations affects the amount of titers waned, and consequentially, the probability of falsely testing negative at post-interval bleed. Figure S2e illustrates this nuance under parameter estimates from Salje, 2018.

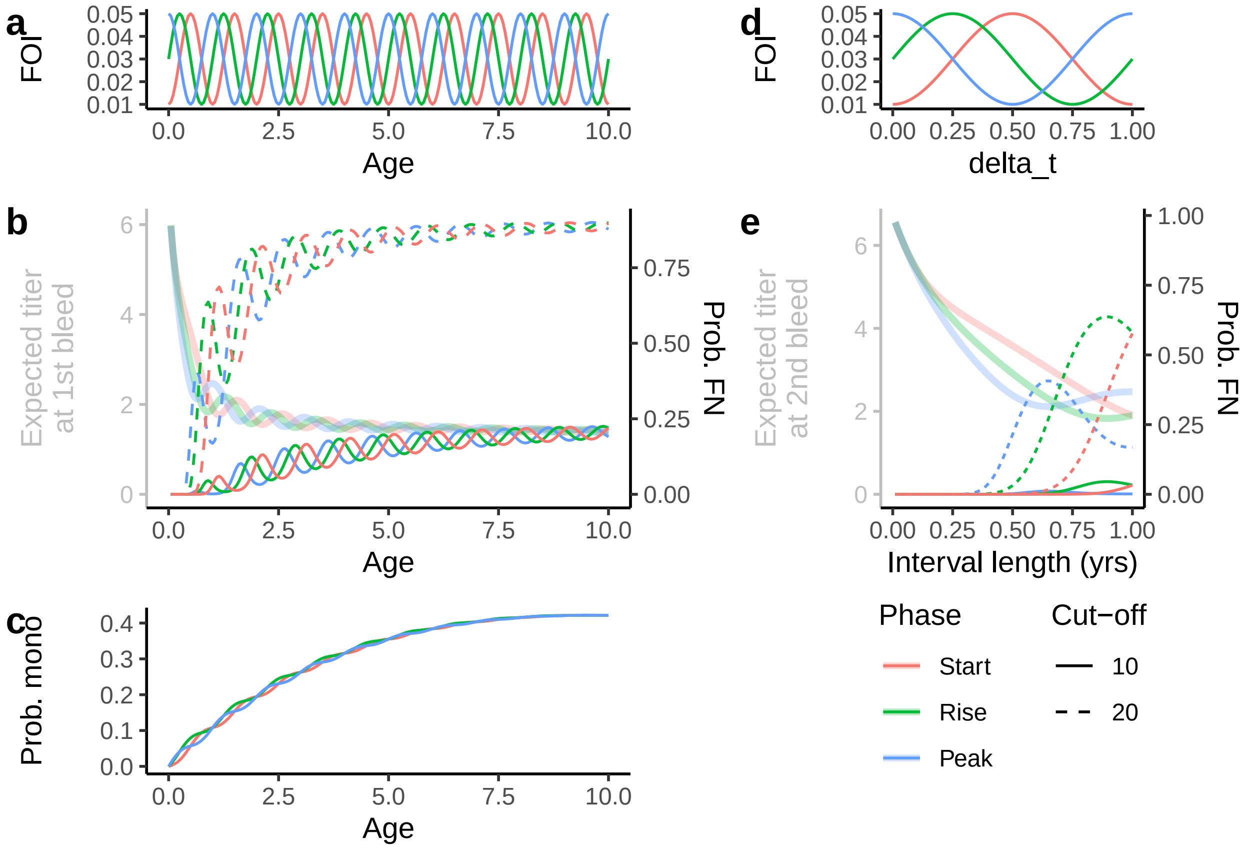


Figure 2: Effects of timings of bleeds in relation to seasonality illustrated using antibody kinetics estimates from Salje, 2018. a) Per-serotype seasonal force of infection (FOI) in age. b) Expected pre-interval titer of a monotypically infected individual (tinted) and the corresponding probability of falsely testing negative given the titers at positivity threshold of 10 (solid) and 20 (dashed). c) Probability that the individual was monotypically infected. d) Relationship between per-serotype seasonal force of infection (FOI) and years since pre-interval bleed. e) Expected post-interval titer of individuals that acquired their first infection during the interval (tinted) and the corresponding probability of falsely testing negative given the titers at positivity threshold of 10 (solid) and 20 (dashed).
